# Case-Control Matching Erodes Feature Discriminability for AI-driven Sepsis Prediction in ICUs: A Retrospective Cohort Study

**DOI:** 10.1101/2025.06.26.25330281

**Authors:** Sophia F. Ehlers, Fanny Tranchellini, Youssef Farag, Tim Hahn, Catherine Jutzeler, Lakmal Meegahapola

**Affiliations:** Department of Medicine, University of Mü nster, Mü nster, Germany; Department of Health Sciences and Technology, ETH Zurich, Zurich, Switzerland; Swiss Institute for Bioinformatics, Lausanne, Switzerland

## Abstract

**Background:** Sepsis remains a leading cause of intensive care unit (ICU) mortality worldwide, and early detection is essential for improving survival through timely interventions. While machine learning holds promise for early sepsis detection by leveraging the multimodal time series data prevalent in ICUs, the field faces significant methodological challenges—such as class imbalance and temporal misalignment. These issues have spurred growing interest in case-control matching strategies. However, there is a lack of systematic evaluation of how commonly used case-control matching methods affect model performance and introduce potential biases, highlighting a critical gap in current knowledge.

**Methods:** We investigated how matched training data affect both the performance of various machine learning architectures and the predictive importance of individual clinical features. Three harmonized large-scale ICU cohorts were used: the high time-resolution ICU dataset (HiRID) from Bern University Hospital, Switzerland (29,698 stays, 2008–2019), with a sepsis prevalence of 6.3%; the Medical Information Mart for Intensive Care (MIMIC-IV) database from Beth Israel Deaconess Medical Center, USA (63,425 stays, 2008–2019), with a sepsis prevalence of 5.2%; and the eICU Collaborative Research Database from 208 US hospitals (123,413 stays, 2014–2015), with a sepsis prevalence of 4.6%. Each dataset included hourly observations. both with and without incorporating demographic variables, and compared the results to unmatched and undersampled cohorts with equivalent ratios. To evaluate how matching strategies influenced feature relevance, we applied a Linear Mixed Effects Model (LMEM) to assess changes in the predictive significance of individual features. Finally, we trained machine learning models—including random forests and gradient-boosted trees—and evaluated their performance on the original test sets using the AUROC and normalized AUPRC metrics.

**Findings:** The LMEM results demonstrated that case-control matching led to a substantial reduction in the number of features identified as significant, based on multiple testing–corrected p-values, across all three cohorts. Specifically, the number of significant features declined from 35–43 in the original datasets to 24–29 as the matching ratio increased from 1:10 to 1:2. In the machine learning experiments, models trained on undersampled data achieved strong performance, with AUROC values exceeding 0.90 and normalized AUPRC scores above 41. In contrast, models trained on the original (imbalanced) datasets performed robustly (AUROC up to 0.82, normalized AUPRC up to 4.2), whereas those trained on matched datasets showed significant performance degradation, with AUROC values below 0.50 and normalized AUPRC scores falling below baseline. These patterns were consistent across all three cohorts and a wide range of evaluation setups.

**Interpretation:** This multi-cohort analysis highlights a methodological paradox: while case-control matching can mitigate class imbalance and help disentangle sepsis-specific patterns from general ICU trajectories, overly strict matching criteria may significantly impair predictive performance. These findings underscore the need for more nuanced matching strategies that balance bias reduction with the preservation of critical clinical signals—ultimately enhancing the reliability and real-world applicability of sepsis prediction models.

**Funding:** This project was supported by grant #902 of the Strategic Focus Area “Personalized Health and Related Technologies (PHRT)” of the ETH Domain and Young Investigator Grant of the Novartis Foundation for Medical-Biological Research.

**Research in Context:** *Evidence before this study:* Case-control matching has long been used in epidemiological research to mitigate confounders and address class imbalance. In recent years, AI-driven sepsis prediction studies have adopted similar strategies to improve data set balance and reduce temporal bias. However, the direct impact of case-control matching on machine learning (ML) performance in sepsis prediction has not been systematically evaluated. Although prior work demonstrated the promise of early detection of sepsis using observational data without explicit matching strategies^1,2^, studies that implemented stringent matching occasionally reported deteriorations in model performance^3,4^. A recent review also advocated the use of case-control temporal alignment to avoid temporal biases^2^. This discrepancy highlights a critical knowledge gap about how various matching protocols influence ML outcomes in sepsis, an inherently heterogeneous condition shaped by both clinical and temporal complexities.

*Added value of this study:* We conducted a comprehensive multi-cohort investigation of absolute-onset case-control matching (with and without demographic criteria) for sepsis prediction models, using three large, harmonized ICU cohorts (HiRID, MIMIC-IV, and eICU). By comparing matched, aligned, and undersampled training cohorts across multiple machine learning architectures and temporal window configurations, our experiments show how matching affects both overall model performance and the discriminability of individual features. We demonstrated that while case-control matching effectively reduces class imbalance, it can also diminish key sepsis-related signals and substantially reduce predictive accuracy below baseline. These insights shed light on the nuanced interaction between epidemiological design strategies and ML-based sepsis prediction in ICU settings.

*Implications of all the available evidence:* Although case-control matching is often viewed as a reliableethod to minimize bias and improve dadata setbalance, our findings indicate that it may inadvertently obscure clinically significant signals critical for accurate prediction of sepsis.is. In real-world clinical environments where timely and precise diagnosis is essential, researchers and clinicians should carefully consider the trade-offs imposed by strict matching protocols, especially when aligning data from large multicenter cohorts. To facilitate transparent model validation, we recommend using absolute-onset matching as a comparative benchmark to prevent artificially inflated performance metrics. Alternatively, temporal dependencies related to admission to the ICU or matched onset should be factored into the model evaluation to avoid trivial classification driven by inadequate handling of within-stay dynamics. Lastly, relying solely on AUROC and AUPRC as performance metrics risks overlooking other clinically relevant outcomes, emphasizing the need for broader evaluation frameworks that align more closely with real-world clinical decision making. This study encourages the research community to refine and reassess conventional matching methods, striving for clinically informed strategies that reduce confounding while retaining essential sepsis-specific signals for robust and generalizable predictive models.

## 1 Introduction

Sepsis, a life-threatening condition characterized by organ dysfunction resulting from a dysregulated host response to infection, remains a major contributor to mortality in intensive care units (ICUs) worldwide^2^. Despite advances in antimicrobial therapies and critical care practices, in-hospital mortality rates for sepsis continue to exceed 20%^5^. Early detection and timely intervention are critical, as prompt administration of appropriate antibiotics is among the most effective strategies for improving surviva^6^. Notably, each hour of delay in antibiotic treatment is associated with a significant increase in mortality risk. This time-critical nature of sepsis management has spurred significant interest in predictive tools capable of alerting clinicians to early signs of deterioration—often before overt clinical symptoms emerge^7,8^. The application of artificial intelligence (AI), in particular machine learning (ML), has emerged as a promising avenue for advancing sepsis prediction in critical care settings. ML models, including deep neural networks and robust ensemble tree methods, are capable of analyzing the high-dimensional, multimodal data found in electronic health records (EHRs) to identify subtle patterns that may precede clinical recognition of sepsis^9^.Interestingly, while deep learning techniques have made considerable strides in ICU prediction tasks, recent studies suggest that ensemble methods such as boosting algorithms and random forests often outperform deep learning models in practical applications^10–13^. Moreover, these models have demonstrated the ability to predict sepsis onset as early as 12–24 hours in advance—and in some cases, even earlier—prior to clinical recognition by healthcare providers. ^14,15^. Such early warning systems could enable timely clinical interventions, illustrating the substantial impact AI can have in enhancing decision-making and outcomes in acute care settings.

Despite recent advances, translating AI models for sepsis prediction into reliable and impactful bedside tools remains a complex challenge. A fundamental obstacle lies in the inherent heterogeneity of critical care environments, which differ substantially across hospitals and regions due to variations in patient populations, data collection frequencies, and clinical protocols^16^. Additionally, the definition of sepsis has evolved over time, with the current Sepsis-3 criteria defining it as an increase of over ≥2 points in the Sequential Organ Failure Assessment (SOFA) score in the presence of suspected infection^17^. However, the practical application of these criteria remains inconsistent, potentially introducing noise and bias in outcome labeling. One of the most significant challenges, even within a single cohort, is the pronounced class imbalance inherent in ICU data, where sepsis cases are vastly outnumbered by non-septic admissions. This imbalance is often compounded by differences in ICU length of stay and clinical trajectories between septic and non-septic patients, increasing the risk that models will learn spurious or non-causal associations^2^. For example, a model might erroneously associate longer ICU stays with sepsis, simply because septic patients are more likely to remain hospitalized for extended periods. Similarly, it may overemphasize age-related risk without capturing the underlying physiological mechanisms. These factors collectively complicate the development of robust, generalizable AI models for early sepsis detection.

One proposed methodological approach to address the challenges of class imbalance and temporal and demographic biases in sepsis prediction is case-control matching, an epidemiological technique that has been adapted for training ML models. In this approach, each identified sepsis case is paired with one or more non-sepsis control patients who exhibit similar baseline characteristics, such as age and pre-existing conditions, and who have a comparable duration of observation within the ICU^2^. By selectively sampling control patients to create a cohort that mirrors the sepsis cases in terms of key confounding variables and the alignment of the time period at risk, matching aims to generate a more balanced dataset. The theoretical advantage of this technique is that it should facilitate the ability of an algorithm to discern patterns specifically related to sepsis by mitigating obvious sources of bias, such as disparities in the length of hospital stays. For example, matching based on the time of sepsis onset after ICU admission, also known as “absolute onset” matching, ensures that a control patient was still present in the ICU at the time when the matched sepsis patient developed the condition, thereby preventing a trivial distinction based solely on the duration of their stay, and its’ effect on the captured features^1,2,18^. The application of case-control matching has become increasingly common in recent sepsis studies seeking to address these inherent data complexities. Beyond temporal alignment, case-control matching offers opportunities to reduce confounding factors and balance distribution shifts of key covariates between cases and controls^19^. For instance, epidemiological research frequently employs case-control matching algorithms based on key patient characteristics, such as demographic features including sex and age^19,20^. This approach is potentially crucial because older patients generally face higher risks for numerous pathologies and clinical complications, including sepsis. Failure to balance training data with respect to age might yield biased outcomes, with prediction models potentially identifying age rather than disease onset. On the contrary, a model should not predict sepsis just because of the age, but should also learn underlying pathophysiologies related to sepsis, beyond age^21^. However, it is important to recognize that matching can introduce selection bias, and the efficiency gains from matching are context-dependent and not universally guaranteed^21^. However, this approach has not consistently enhanced model robustness, as previous research has documented performance degradation in MGP-RNN models following case-control alignment^3,4^. This finding could imply that overly rigid matching protocols might eliminate valuable variability within the data, leading to the creation of an artificial patient cohort that, while statistically more homogeneous, is actually more challenging for an AI model to learn from effectively. In essence, the very process intended to increase precision in the selection of the study cohort could paradoxically reduce the discriminability of the relevant clinical features. These considerations have generated conflicting perspectives regarding the merits, necessity, and limitations of case-control matching and temporal alignment in sepsis prediction, despite the absence of structured, comprehensive examinations of this topic. Current literature lacks standardized reporting procedures for these methodological aspects, and significant knowledge gaps persist concerning the interaction between onset matching and various machine learning architectures.

This study aims to address this critical methodological challenge through a systematic analysis across multiple large-scale, diverse, and harmonized ICU cohorts. We hypothesize that while case-control matching can help control for confounding variables, its stringent application may inadvertently attenuate the predictive signal of clinically relevant features, ultimately compromising model performance. To investigate this, we conduct a comprehensive evaluation of absolute-onset case-control matching—applied with and without demographic feature alignment—and its impact on machine learning–based sepsis prediction. Our objectives are twofold. First, we assess how case-control matching influences the statistical significance and predictive contribution of individual clinical features, using rigorous feature-level analysis. Second, we evaluate how varying matching ratios affect the performance of machine learning models, specifically tree-based and gradient-boosted architectures. We compare models trained on matched cohorts to those trained on unmatched and undersampled datasets with equivalent case-to-control ratios, thereby isolating the effects of the matching strategy itself. Through this investigation, we aim to deepen our understanding of the complex interplay between epidemiological matching techniques and machine learning methods, and to provide guidance for the design of more robust and generalizable predictive models for early sepsis detection.

## 2 Methods

### 2.1 Harmonizing Datasets & Cohort Selection

This study used YAIB pipeline^22^ and RiCU^23^ to harmonize dataset and perform cohort selection. YAIB is a flexible multi-center framework designed for conducting reproducible and comparable machine learning experiments in intensive care settings. Our analysis focused specifically on three cohorts, that are natively supported by YAIB: the High Time Resolution ICU Dataset (HiRID)^12,24^, the Medical Information Mart for Intensive Care IV (MIMIC-IV)^25,26^, and the eICU Collaborative Research Database (eICU)^27,28^. More detailed provided in Table 1. The YAIB workflow initially focused on creating compatible datasets through unit harmonization, artifact removal, and variable mapping. Clinical concepts were extracted using ricu^23^, an R package specifically designed to transform diverse datasets into a common semantically interoperable format. Following the extraction of a Sepsis Cohort and relevant variables, the pipeline facilitated data pre-processing and task definition, allowing us to extract harmonized cohorts from the three diverse databases. Moreover, the harmonized data structure comprised 48 dynamic features, including 7 vital signs, 39 laboratory tests, urine output, and FiO2. To ensure consistency across datasets, data frequency was harmonized to hourly intervals, as MIMIC-IV only provides hourly data input, while eICU and HiRID offer data entries at least every 5 minutes. The maximum duration of data collection was set at 7 days per patient.

**Table 1.** Details of ICU datasets HiRID, MIMIC-IV and eICU. These datasets were used for the YAIB harmonization workflow^22^. Each dataset is publicly available and can be accessed via Physionet after completing a credentialing procedure.

| Dataset |  | HiRID | MIMIC-IV | eICU |
| --- | --- | --- | --- | --- |
| Originally published |  | 2020<br>(Hyland, 2020)<sup>12</sup> | 2020<br>(Johnson, 2020)<sup>25</sup> | 2017<br>(Pollard, 2018)<sup>27</sup> |
| Version |  | v1.1.1 | v2.0 | v2.0 |
| Years of Data Collection |  | 2011-2016 | 2008-2019 | 2014-2015 |
| Origin |  | Switzerland | USA | USA |
| Frequency (time-series) |  | 2/5 minutes | 1 hour | 5 minutes |
| Number of patients |  | 33,905 | 53,090 | 166,916 |
| Number of ICU stays |  | 32,338 | 75,652 | 182,774 |
| Admission type | Medical | – | 49,217 (65%) | 134,632 (79%) |
|  | Surgical | – | 25,674 (34%) | 51,909 (19%) |
|  | Other | – | 761 (1%) | 4,702 (3%) |
| Age at admission (median [IQR]) |  | 65 [55, 75] | 65 [53, 76] | 65 [53, 76] |
| Gender | Male | 20,796 (64%) | 42,153 (56%) | 98,834 (54%) |
|  | Female | 11,542 (36%) | 33,499 (44%) | 83,940 (46%) |
| Race | White | – | 51,575 (76%) | 140,958 (78%) |
|  | Black | – | 8,223 (12%) | 19,887 (11%) |
|  | Asian | – | 2,225 (3%) | 5,008 (3%) |
|  | Other | – | 5,514 (8%) | 16,978 (9%) |

#### 2.1.1 Cohort Selection: Exclusion Criteria

For Sepsis Cohort selection, several exclusion criteria were applied in accordance with clinical definitions^22^. To ensure high data quality, we excluded patients with invalid admission or discharge times, ICU stays shorter than six hours, fewer than four hourly clinical measurements, or more than 12 consecutive hours without any clinical data. Additionally, we excluded patients under 18 years of age at admission and any stays from hospitals that did not report at least one sepsis case, to ensure sufficient data for defining the outcome. We also excluded cases where disease onset occurred outside the ICU or within the first six hours of ICU admission, enabling six-hour back-labeling of sepsis onset and supporting early prediction.

#### 2.1.2 Sepsis Definition

Sepsis onset was defined as the first instance of organ dysfunction, characterized by a ≥2 point increase in the SOFA score within 24 hours, occurring within 48 hours before and 24 hours after suspected infection^17,29^. Suspected infection was identified based on the concurrent administration of antibiotics and the culture of body fluids, with cultures collected within 24 hours after antibiotic initiation or antibiotics started within 72 hours after culture collection. For HiRID and eICU, which lack microbiological data, suspicion of infection was determined solely by antibiotic administration. To exclude prophylactic use, antibiotics had to be administered for at least 3 days or for the entire ICU stay^30^. A “True” (Sepsis) label was assigned starting 6 hours before the determined onset, framing the task as a prediction problem, with a cutoff 12 hours after the onset, defining this as the onset period. This methodology yielded sepsis prevalences of 6.3% for HiRID, 5.2% for MIMIC-IV, and 4.6% for eICU (Table 2).

**Table 2.**
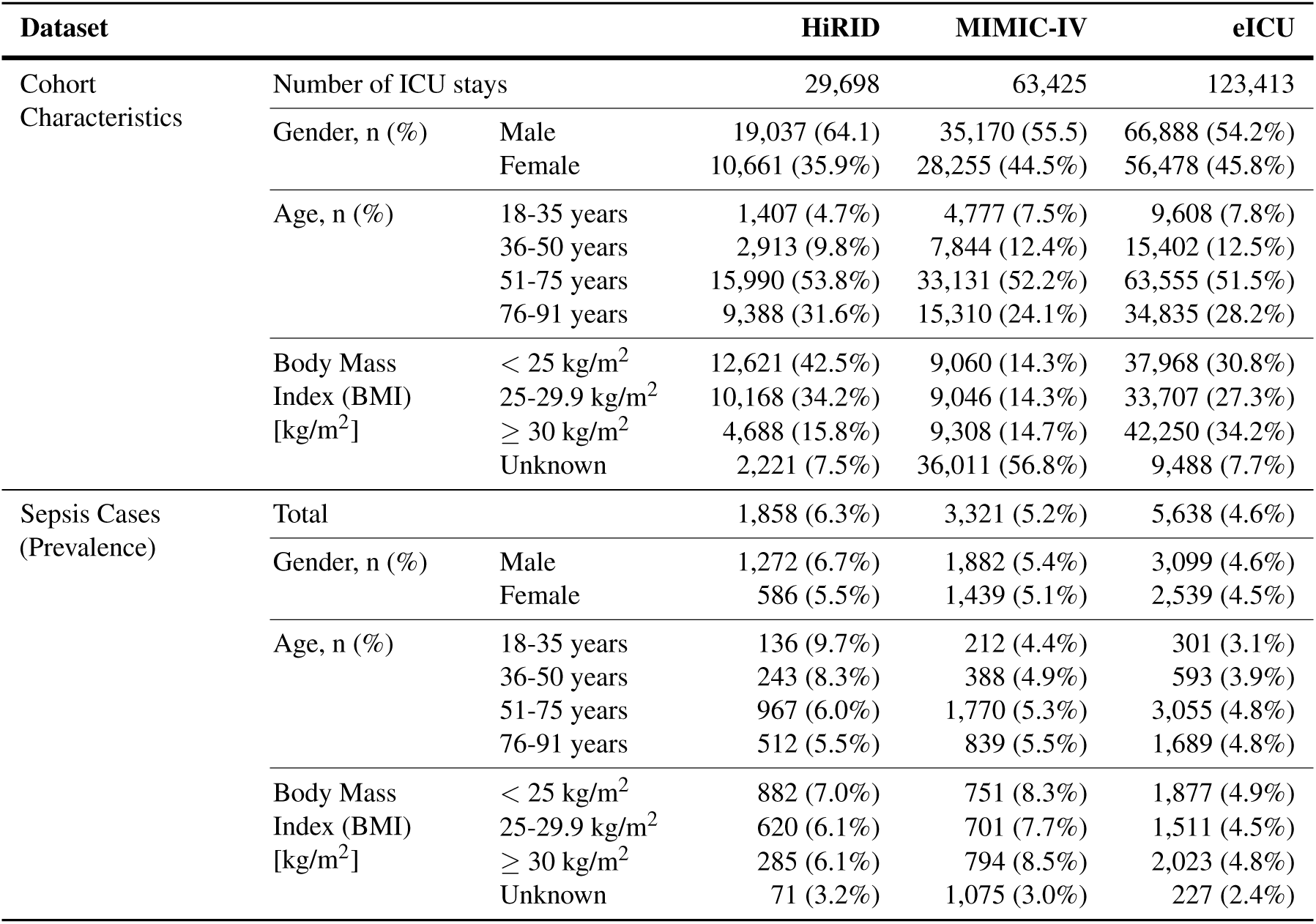
Details of harmonized ICU cohorts HiRID, MIMIC-IV and eICU. Cohort characteristics and sepsis prevalence were analyzed following harmonization, with sepsis task and cohort definitions established according to the YAIB framework^22^.

### 2.2 Data Preprocessing Pipeline

The preprocessing methodology followed a systematic approach to prepare intensive care unit data for machine learning analysis.

#### 2.2.1 Clinical Domain-Informed Outlier Handling

Clinical measurements in intensive care settings are susceptible to errors arising from equipment malfunction or mishandling. The interpretation of vital signs and laboratory test results necessitates comprehensive evaluation of the clinical situation. Our harmonized datasets exhibited a considerable number of outliers across dynamic features. Each feature required careful evaluation to distinguish between dismissible (physiologically implausible) outliers and values indicative of severe patient conditions^31^. We implemented a comprehensive outlier detection approach grounded in clinical knowledge. For each physiological parameter, we defined both normal physiological ranges and extreme permissible values as given in Table 3. These permissible limits were expanded by 5% of the permissible range in both directions and values outside these expanded ranges were converted to missing values (NaN) to preserve the temporal structure of the data.

The dataset was divided into training (90%) and testing (10%) sets using sklearn’s GroupShuffleSplit, ensuring that all measurements from individual patient stays remained together. This approach prevented data leakage and maintained the temporal integrity of patient trajectories, which also taking into account admission time to the ICU, to obtain a temporal split. The resulting split preserved comparable frequencies of Sepsis Cases and Sepsis Labels between training and testing sets (Table 5).

We applied the Standard-Scaler from the Scikit-learn^32^ library to transform both dynamic clinical measurements and static numeric features (age, weight, height). Each feature was standardized to zero mean and unit variance to facilitate model training, while sex as a categorical variable was encoded as −1/1. To prevent data leakage, mean and variance calculations were derived exclusively from the training set, and then applied to the testing set with learned mean and variance.

The handling of missing values proved particularly crucial given the nature of clinical time series data. Laboratory results frequently exhibited over 90% missing data, primarily due to daily measurement frequencies being represented in hourly data input. Comparing percentage and pattern of missing data across datasets and groups (sepsis, non-sepsis) did not reveal major differences that would prevent applying the same missing value strategy for all datasets and groups (Figure 2A, Figure 2B). We developed a two-pronged approach to address missing values. First, we created binary missingness indicators for each feature to explicitly capture missing value patterns. Second, we implemented hierarchical imputation, beginning with static features (demographics) using patient-specific forward/backward fill. For stay_ids lacking values, we assigned the global mean, and for gender, the mode (male). Dynamic features were imputed using a combination of forward fill within each patient stay and global mean imputation as a fallback, with global means calculated from the training set to prevent data leakage. This strategy was designed to be applicable in realistic clinical settings where new patients are admitted to the ICU.

### 2.3 Case-Control Matching

#### 2.3.1 Matching Strategy and Criteria

Our overall preprocessing pipeline (see Figure 1A) begins with domain-informed outlier handling and proceeds through three distinct case-control balancing strategies: (Pathway 1) *undersampling* of random non-sepsis stays, (Pathway 2) *alignment* of randomly sampled non-sepsis stays using absolute onset matching for temporal window alignment, and (Pathway 3) *matching* with demographic criteria (*age*, *height*, *weight*, and *sex*) while also performing the same temporal alignment steps. These three strategies are then followed by data splitting, standardization, and missing value imputation, that were mentioned earlier. To illustrate the effect of these strategies, Figure 1C schematically contrasts absolute onset matching (with and without demographic variables) with undersampling, the latter lacking temporal alignment. To mitigate the class imbalance inherent in sepsis prediction, we employ the *matching* strategy (pathway 3 above) to ensure balanced representation of sepsis and non-sepsis cases while preserving clinical plausibility. In particular, *absolute onset matching* is used to prevent biases arising from systematically different length-of-stay distributions of sepsis vs. control groups^1,2^. Controls must have a monitoring duration at least as long as the matched case’s pre-onset period, and data collected beyond 12 h post-matched absolute onset are excluded from further analysis. For demographic matching, primary matching criteria are: age within ±5 years, height within ±10 cm, weight within ±10 kg, and exact matching for sex. Accounting for age and sex is standard practice in clinical studies^33,34^, while weight and height (and thus approximate body-mass index) are included due to their reported effects on sepsis outcomes^35^. An ablation analysis was performed to investigate the contribution of these demographic features by removing them for both model training and testing. Across all three cohorts, the distribution of static features showed only minor variations across cases/controls and by gender (Figure 3), justifying a consistent approach.

**Figure 1.**
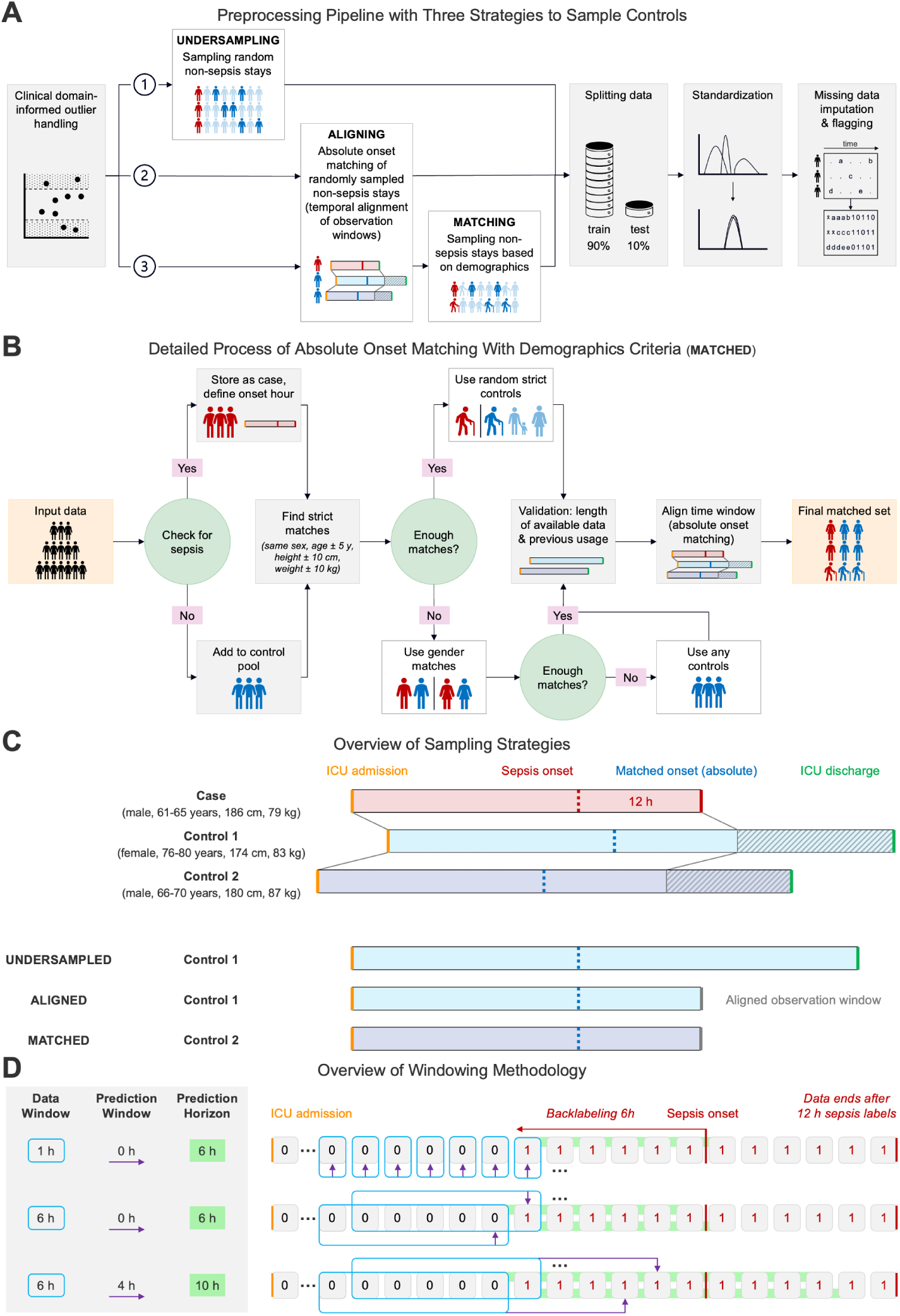
Case-control matching and windowing methodology. **A)** Preprocessing pipeline comprising outlier management, three case-control balancing strategies (1. undersampling, 2. alignment, and 3. matching – incorporating temporal alignment with demographics-based matching), followed by data splitting, standardization, and missing value imputation. **B)** Detailed process of absolute onset matching using demographics (”matched”, A3). Sepsis cases are identified and time to onset determined. Controls are selected using a hierarchical matching approach: primary matching within predefined demographic parameter thresholds, secondary matching based on biological sex, and tertiary random assignment when other matching criteria could not be satisfied. Each control subject is utilized only once, with temporal data windows aligned between matched case-control pairs. **C)** Schematic illustration of absolute onset matching implementation with and without demographic variables, contrasted with undersampling methodology lacking temporal alignment. Mentioned demographic information are hypothetical, done for this illustration. *Absolute* refers to the absolute time since ICU admission. Controls must have monitoring duration equal to or exceeding their matched cases. All data collected beyond 12 hours post-matched absolute onset are excluded from analysis. **D)** Methodological Overview of temporal windowing with implementation of three data-prediction-window pairings: 1-hour/0-hours, 6-hours/0-hours, and 6-hours/4-hours. Positive sepsis labels are initiated six hours before officially defined sepsis onset (”backlabeling”) and persist for 12 hours total, penalizing late false-negative predictions during model training. This approach yields a 6-hour prediction horizon when no prediction window (0 hours**_5_**)**_/3_**is^6^ applied. Predictive assessments occur at hourly intervals.

**Figure 2.**
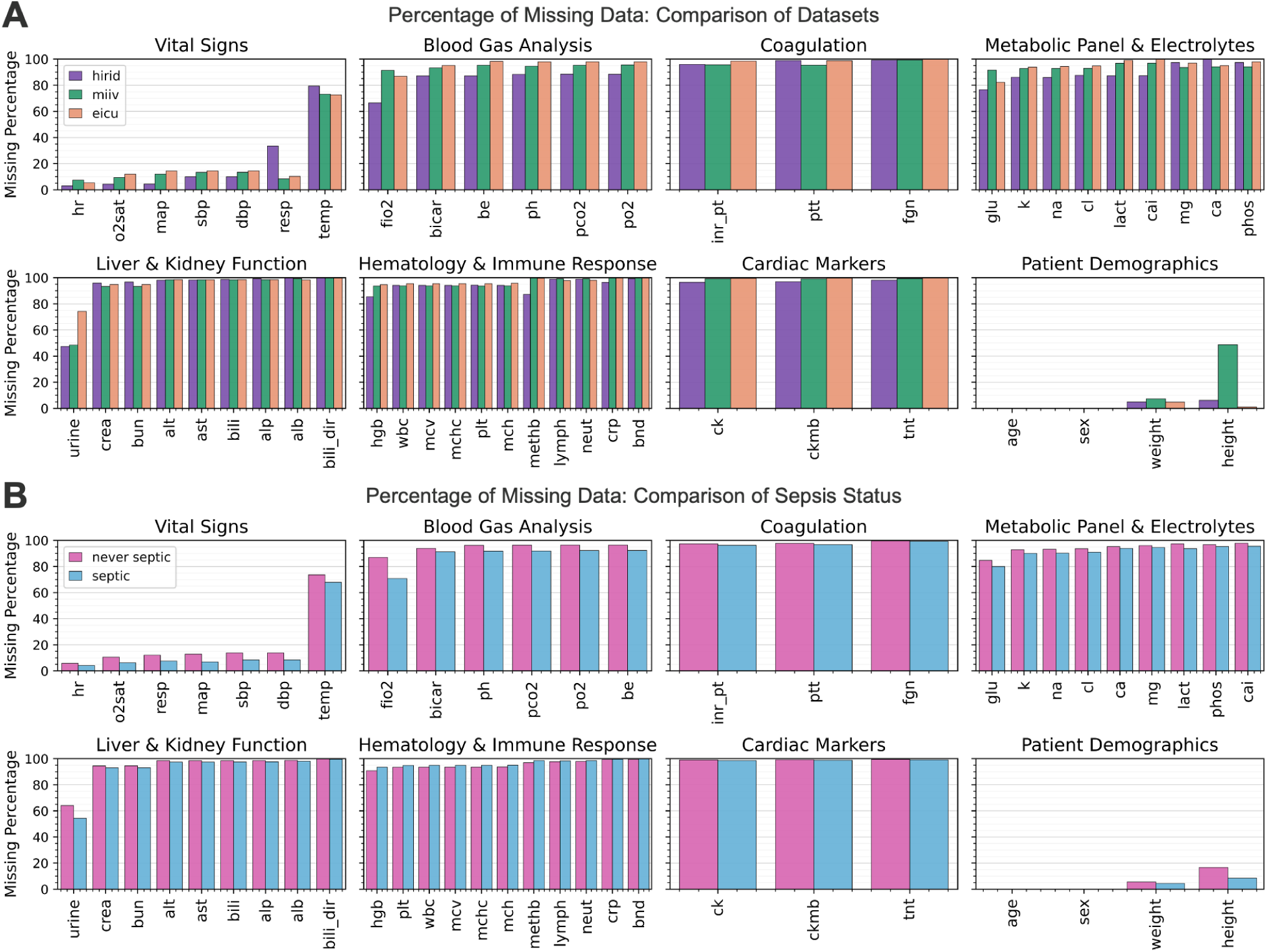
Percentage of missing data across features, datasets and sepsis status. Clinical time series data inherently exhibits substantial missingness due to the sparse and irregular sampling intervals employed during data collection. The multi-modal nature of intensive care unit datasets results in heterogeneous patterns of missing values across measurement types. Vital sign parameters are recorded at higher temporal frequencies compared to laboratory values, creating differential missingness profiles. Conversely, static patient characteristics such as demographic variables demonstrate near-complete data integrity with minimal missing entries. Percentages of missing data across all 48 dynamic and 4 static features in the harmonized datasets, stratified by dataset **(A)** and sepsis status (patients that are never septic and patients with confirmed or incipient sepsis) **(B)**. position of each measurement. In instances where strict demographic matching cannot be met, we implement a two-tier fallback system to avoid losing sepsis cases^36^: (i) match on sex only, then (ii) allow any remaining controls with sufficient observation length. Each control is used only once (matching without replacement), thereby reducing selection bias. To investigate different effective sepsis prevalences, three target ratios of controls per sepsis case (10, 5, and 2) are sampled. Although this increases the proportion of sepsis in the final datasets relative to typical ICU prevalence, these values remain within ranges reported in multiple sepsis studies^2^. Matching was performed consistently all three cohorts, leading to decreasing overall cohort size as the sepsis ratio increased (Table 5, Figure 12). The same ratios were also applied during undersampling to provide a point of comparison, ensuring that performance increases or decreases are not merely due to changes in the number of patients within the cohorts, but rather the matching strategy itself.

**Figure 3.**
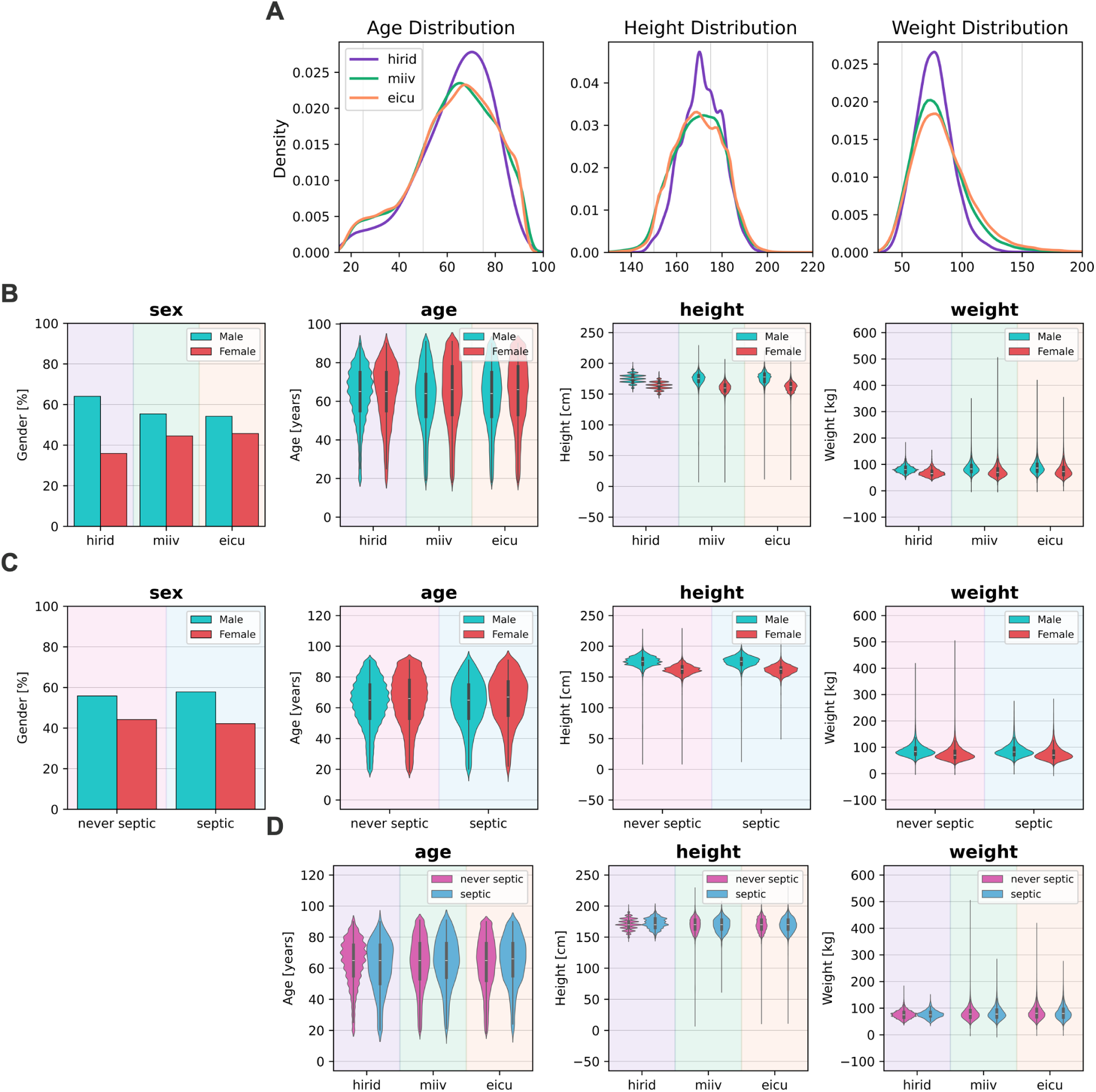
Distribution analysis of static features. **A)** Age, height, and weight distributions across three critical care datasets: HiRID, MIMIC-IV, and eICU. **B)** Comparative analysis of static clinical variables stratified by biological sex across the three datasets. **C)** Static clinical variables stratified by biological sex with comparative analysis between patients without sepsis (never septic) and those with confirmed or incipient sepsis (septic). **D)** Comparative analysis of static clinical variables stratified by age categories across the three critical care datasets.

#### 2.3.2 Matching Algorithm Implementation

The matching process is summarized in Figure 1B (“matched”, A3) and involves first identifying sepsis cases and their onset times. We then create a pool of non-sepsis controls that satisfy (i) the required demographic thresholds and (ii) the temporal alignment requirement (i.e., sufficient observation length to match each sepsis case). The *absolute* time since ICU admission is used to align each control’s time series with the case’s onset hour; a “time_to_onset” variable is introduced to track the relative

### 2.4 Data Windowing

#### 2.4.1 Window Design and Components

To transform continuous patient time series into structured input for machine learning models, we implemented a sliding window approach, sometimes referred to as “online training,” which creates fixed-length data windows with varying prediction windows^2,30,37^ (Figure 1D). Windowing was performed after preprocessing and potential case-control matching steps. This methodology represents a standard approach for ICU prediction tasks based on structured time-series data, enabling models to identify temporal trends and patterns while evaluating their predictive capabilities across different time horizons. The sliding window technique offers greater sophistication than simply utilizing data from the minimum time interval (one hour) for sepsis classification. Notably, due to the six-hour backlabeling window applied during task definition, even this simpler implementation constitutes a six-hour prediction horizon^18,30,38,39^. We established a step length of 1 for the sliding window approach, maximizing the number of windows generated per stay_id. Only windows containing complete data and prediction windows were created, which resulted in fewer available windows with increasing window sizes.

The data window (observation window) was designed with a standardized length of 6 hours, which aligns with the YAIB exclusion criteria stipulating that patients must remain in the ICU for a minimum of six hours^22^. This parameter ensures that each stay_id generates at least one data window. The data window encompasses all dynamic clinical measurements and laboratory values while preserving the temporal sequence of observations. Static patient characteristics (age, sex, height, weight) were replicated across all windows to maintain consistency throughout the analysis. A gap between the data window and the targeted prediction time point is defined as a prediction window^40^, while the prediction horizon comprises the sum of the prediction window and the backlabeling window. We implemented two prediction window configurations: first, no prediction window, utilizing the sepsis label from the final hour of the data window, resulting in a six-hour prediction horizon due to backlabeling; second, a four-hour prediction window, yielding a ten-hour prediction horizon to assess model performance for earlier prediction. This represents a prediction horizon that, while noted as challenging, has been demonstrated to be consistently achievable in previous research^41^. Earlier prediction capability would provide significant clinical benefit, allowing healthcare providers additional time to evaluate patient conditions and initiate treatment promptly. This consideration is particularly relevant as current machine learning models demonstrate higher accuracy in short-term rather than long-term sepsis prediction^42^.

#### 2.4.2 Implementation and Validation

The sliding window implementation processes each patient stay sequentially. For each possible starting position, a six-hour data window is created, with the values of all features being stacked (e.g., feature1_win0 until feature1_win5) and combined with the static patient information. The resulting data structure represents each window as a single row, containing both the stacked features and their corresponding label extracted from the appropriate prediction point. The validation process ensures data quality and temporal consistency through several critical requirements. Patient stays must be sufficiently long to accommodate both the data window and prediction window, with a step-length of 1 hour implemented to prevent data loss. The temporal integrity of measurements within each window is preserved, maintaining the original sequence and relationships of the clinical data. Furthermore, careful attention is paid to the alignment of outcome labels with their corresponding windows, ensuring accurate temporal relationships based on the designated prediction horizon. This comprehensive window creation methodology was applied uniformly across all three cohorts, as well as across all case-control matching variants (original, 1:10, 1:5, 1:2), ensuring consistency in the temporal representation of patient data throughout the analysis.

### 2.5 Linear Mixed Effects Models

#### 2.5.1 Model Specification and Fitting

To assess the differences between septic and never-septic patients (cases and controls) while accounting for repeated measurements and individual patient variability, we implemented a Linear Mixed Effects Model (LMEM) analysis framework^43^. LMEMs incorporate both fixed effects, representing population-level parameters, and random effects, capturing variability within groups. For each clinical dynamic variable, we fitted a separate univariate LMEM of the form: *Variable* ∼ *Group* + (1 | *PatientID*) where the dependent variable represents the standardized clinical measurement (dynamic features), Group is a binary indicator (1 for septic cases, 0 for controls), and (1|PatientID) represents random intercepts for each patient stay. Models were fitted using restricted maximum likelihood estimation (REML)^44^. This model structure allows us to account for the repeated measurements of each variable within patients and estimate the fixed effect of sepsis status on each variable separately. Model diagnostics were performed, including assessment of convergence for each model, examination of variance components, and evaluation of model fit through random effects variance and residual analysis.

#### 2.5.2 Statistical Analysis and Implementation

The statistical analysis framework incorporated multiple levels of inference and validation. P-values were adjusted for multiple comparisons using Benjamini-Hochberg FDR correction^45^, and confidence intervals were calculated for fixed effects. Intraclass Correlation Coefficients (ICC)^46^ were computed to assess the proportion of variance explained by patient-level clustering. The LMEM analysis was conducted across multiple dimensions, utilizing training sets that resulted from preprocessing with or without case-control matching and without windowing (Table 5, Figure 12). Models were fitted separately for each ICU dataset (HiRID, MIMIC-IV, eICU) to assess consistency across different data sources. The impact of case-control matching was evaluated by performing analyses on both the original and matched datasets, with multiple matching ratios (1:2, 1:5, 1:10) being considered. For effect size interpretation, coefficient estimates represent the standardized mean difference between cases and controls, with positive coefficients indicating higher values in septic patients. Effects were considered significant at an FDR-adjusted p-value < 0.05. This comprehensive approach allowed us to systematically evaluate the differences between septic and non-septic patients while accounting for the complex structure of intensive care unit data.

### 2.6 Machine Learning Models

#### 2.6.1 Classification Models

For the prediction of sepsis onset, we implemented multiple machine learning approaches, encompassing both traditional ensemble methods and gradient boosting algorithms. Recent studies demonstrated equal or stronger performance of traditional machine learning models as compared to deep learning models for ICU prediction tasks^11–13^. Four distinct classification models were selected based on their specific characteristics relevant to sepsis prediction. Random Forest (RF)^47^ and Balanced Random Forest (bRF)^48^ were implemented using sklearn’s RandomForestClassifier and imblearn’s BalancedRandomForestClassifier respectively. While the standard RF implementation leverages ensemble learning with multiple decision trees maintaining the original class distribution, bRF incorporates automatic adjustments for class imbalance during training, making it particularly suitable for the natural class imbalance in sepsis prediction. Additionally, we employed two gradient boosting frameworks: LightGBM (LGBM)^49^ and XGBoost (XGB)^50^. LGBM utilizes a leaf-wise tree growth strategy and was configured with a binary classification objective. XGBoost, incorporating additional regularization features, was optimized for binary classification tasks using AUC as the evaluation metric during training. To ensure reproducibility and establish baseline performance, all “vanilla” models were implemented with consistent configuration parameters, including a fixed random seed of 42, enabled parallel processing (n_jobs=-1), binary classification objectives where applicable, and default hyperparameters.

#### 2.6.2 Model Optimization with Hyper-Parameter Tuning

For the best performing vanilla models, we optimized our model using a Bayesian optimization approach^51^, utilizing a sequential approach to hyperparameter tuning that efficiently balances exploration and exploitation of the parameter space. The hyperparameter ranges for each model are specified in Table 6. The optimization protocol comprised five initial random points for exploration, followed by 25 sequential optimization iterations. Performance was evaluated using 5-fold cross-validation with AUROC as the optimization metric, and parallel processing was enabled. This optimization process was applied independently to each model and dataset combination, ensuring optimal adaptation to the specific characteristics of each ICU cohort while maintaining computational efficiency.

#### 2.6.3 Evaluation of Model Performance

Model evaluation incorporated two complementary performance metrics: Area Under the Receiver Operating Characteristic Curve (AUROC) and Area Under the Precision-Recall Curve (AUPRC). AUROC, while widely established in sepsis prediction literature and valuable for cross-study comparisons, exhibits limitations in imbalanced datasets due to its equal weighting of false positives and false negatives. An AUROC value of 0.5 represents performance equivalent to random classification, with values above 0.5 indicating increasingly better discrimination. To address the limitations of AUROC in class-imbalanced scenarios, we incorporated AUPRC as an additional evaluation metric, given its enhanced sensitivity to positive class predictions in imbalanced datasets. Since baseline Precision-Recall curves demonstrate dependence on class prevalence, we implemented normalization to enable valid cross-dataset comparisons. A normalized AUPRC (nAUPRC) of 1 reflected baseline performance (random guessing), while values exceeding 1 indicated improved predictive performance compared to baseline. Normalized AUPRC was defined as:

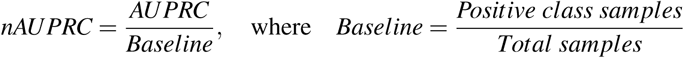

The evaluation framework was designed to be dataset-agnostic, facilitating consistent comparison across the three ICU cohorts, while maintaining the ability to capture dataset-specific performance characteristics.

## 3 Results

### 3.1 Case-Control-Matching Reduces Significant Feature Differences Between Cases and Controls

Our LMEM analysis demonstrated that case-control matching substantially influenced FDR-corrected significance levels across features and datasets (Figure 4). A notable finding was the inverse relationship between sepsis prevalence and the retention of significant features, attributed to decreasing matching ratios (Figure 4A,B).

**Figure 4.**
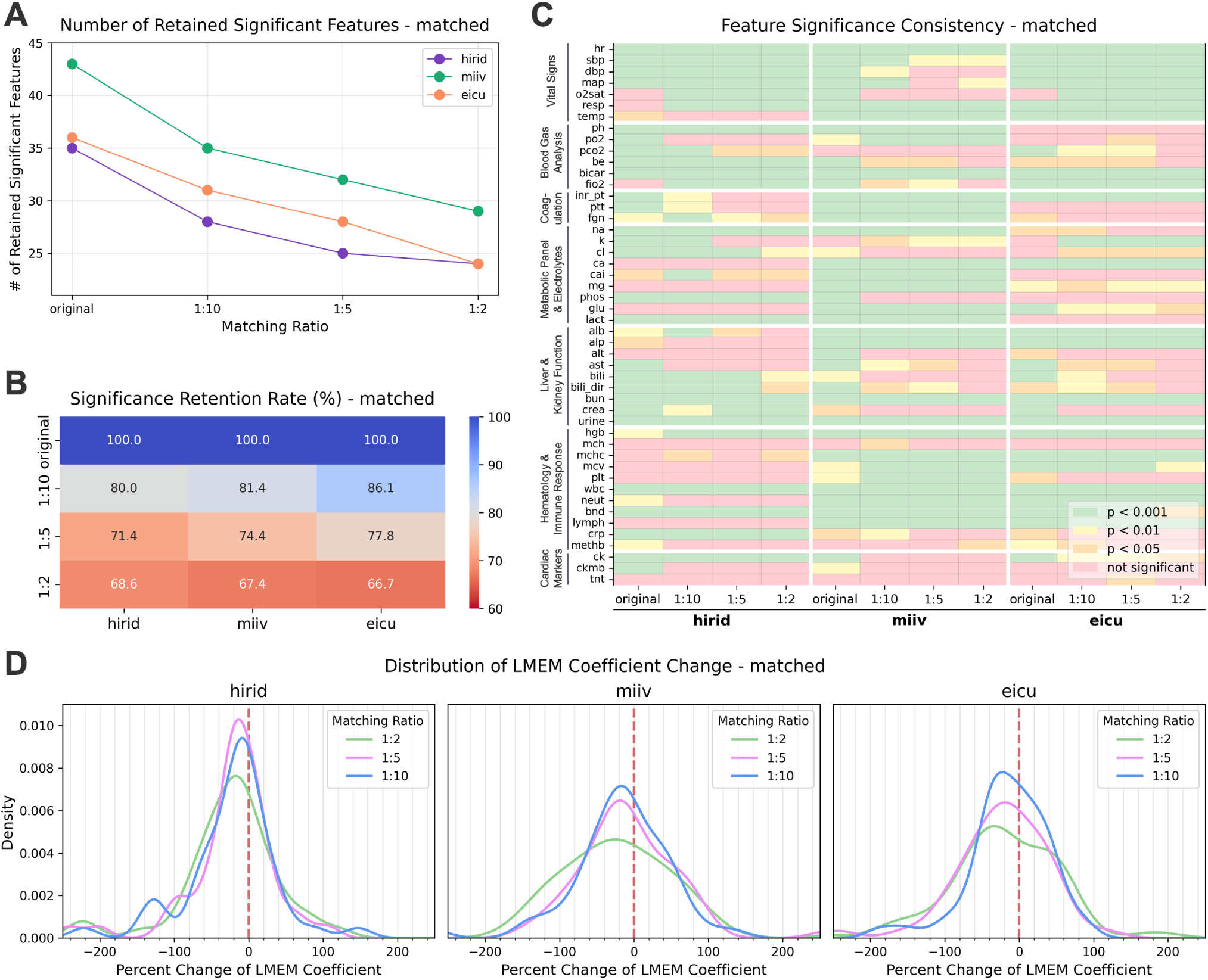
Evaluation of LMEM coefficient alterations and their statistical significance (original vs. matched). **A)** The absolute number of retained significant features in matched training cohorts compared to original training sets demonstrates an inverse relationship with increasing matching ratio. **B)** The relative significance retention rate exhibits comparable patterns across all three ICU datasets. **C)** Analysis of significance levels reveals distinct heterogeneity in feature significance profiles across datasets. **D)** The distribution shift toward negative percent changes in LMEM coefficients indicates reduced discriminative capacity of the collective dynamic features for differentiating between sepsis and non-sepsis cohorts.

**Figure 5.**
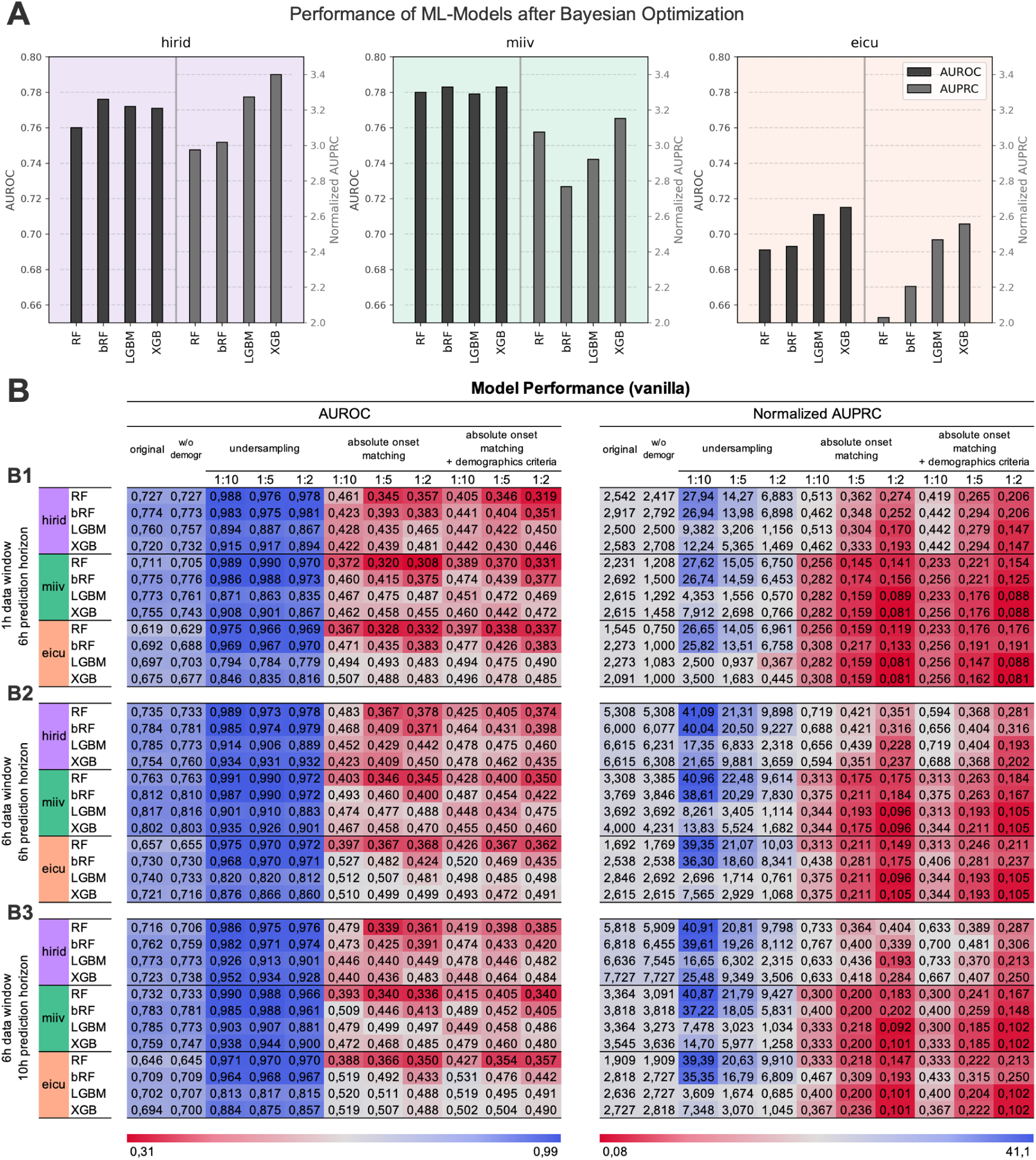
Performance of machine learning models. **A)** AUROC and nAUPRC metrics of machine learning models following Bayesian hyperparameter optimization, trained and evaluated on original datasets. **B)** Comparative analysis of non-optimized (vanilla) machine learning models trained on specified training datasets (original, demographics-excluded, as well as undersampled, aligned, and matched at 1:10/1:5/1:2 ratios each) and evaluated on the original test cohort. Left panels display AUROC metrics while right panels present nAUPRC metrics. This experimental framework was implemented across three distinct temporal configurations: **(B1)** 1-hour data window with 0-hour prediction window, **(B2)** 6-hour data window with 0-hour prediction window, and **(B3)** 6-hour data window with 4-hour prediction window. Models trained on undersampled cohorts demonstrate substantial performance metric improvements, while those trained on absolute-onset matched cohorts—with or without demographic criteria—exhibit performance metrics below baseline levels.

In the HiRID dataset, of the 48 time series variables analyzed, we observed a progressive reduction in significant features: 35 (100.0%) variables showed significant group differences in the original dataset, decreasing to 28 (80.0%) in the 1:10 matched training set, 25 (71.4%) in the 1:5 matched set, and 24 (68.6%) in the 1:2 matched set. The eICU dataset exhibited marginally more significant features initially, with 36 (100.0%) significant features in the original dataset, declining to 31 (86.1%) at 1:10 matching, 28 (77.8%) at 1:5 matching, and 24 (66.7%) at 1:2 matching. While the MIMIC-IV dataset presented the highest absolute number of significant features, it followed a similar declining pattern: 43 (100.0%) significant features in the original dataset, 35 (81.4%) at 1:10 matching, 32 (74.4%) at 1:5 matching, and 29 (67.4%) at 1:2 matching. These consistent patterns across all analyzed datasets suggest that case-control matching similarly affects the discriminative power of features between sepsis and non-sepsis patients.

Across ICU datasets, numerous dynamic features exhibited comparable significance levels (Figure 4C). We established significance for FDR-corrected p-values at the 0.05 threshold and further stratified significant features into three categories: p<0.05, p<0.01, and p<0.001. Vital signs including heart rate, systolic blood pressure, and mean arterial pressure demonstrated significant differences between sepsis and non-sepsis groups consistently across all datasets and matching variants. However, temperature showed reduced predictive value in HiRID, while oxygen saturation reached significance only in the original MIMIC-IV training set and across all matching variants of HiRID and eICU. The varying significance patterns across vital signs highlighted the complexity of sepsis detection using traditional clinical parameters.

Notably, while white blood cell count, a major inflammation marker, maintained significance throughout, C-reactive protein (CRP) showed consistent significance in HiRID variants but demonstrated reduced significance in MIMIC-IV and eICU. Lactate, a key indicator of tissue hypoperfusion and compromised oxygen delivery in septic shock, proved significant across all variants of HiRID and MIMIC-IV, but unexpectedly failed to reach significance in any eICU variant. These findings underscore the variability in the predictive value of established sepsis biomarkers across different clinical settings. Our analysis of organ function markers revealed distinct patterns, with urine output maintaining consistent significance while creatinine lost significance in matched variants of MIMIC-IV and eICU. Bilirubin, a surrogate marker for liver function, similarly showed reduced significance in certain variants of MIMIC-IV and eICU, suggesting potential regional differences in liver function assessment or documentation.

Despite dataset harmonization efforts, substantial differences in feature predictiveness persisted between the analyzed ICU cohorts. The most pronounced disparities were observed between HiRID, a Swiss dataset, and the American datasets MIMIC-IV and eICU (Table 1). These geographical variations in feature significance highlight the importance of considering regional healthcare practices and documentation standards when developing sepsis prediction models.

Our investigation next focused on quantifying the magnitude of LMEM coefficient changes across different matching ratio variants, both for aggregate features and individual parameters. In LMEMs with binary outcomes, coefficients indicate the probability change of outcome occurrence per unit increase in the predictor while accounting for random effects. A coefficient approaching zero suggests a diminished expected effect on the outcome, which can be expressed as a negative Percent Change. The analysis revealed that matched training sets consistently demonstrated a shift toward negative Percent Changes across all ICU datasets and matching-ratio variants (Figure 4D). These findings were derived from the absolute values of LMEM coefficients (Figure 6), revealing distinct patterns across different clinical parameters. Dynamic features associated with vital signs, blood gas analysis, and metabolic panel and electrolytes demonstrated notably larger coefficient values compared to coagulation and cardiac markers. Interestingly, the 95% Confidence Intervals showed considerable variation across datasets, with eICU exhibiting the narrowest intervals and HiRID the widest, potentially attributable to differences in dataset size (Table 1). This variation in confidence intervals underscores the importance of considering dataset characteristics when interpreting the robustness of predictive features.

Moreover, the appendix (Figure 9 and Figure 10) shows the LMEM results comparing the original versus aligned cohorts (Pathway 2 in Figure 1A). The absolute number of retained significant features in aligned training cohorts, compared to the original training sets, displays an inverse relationship with increasing matching ratios. This relationship resembles the results from matched training cohorts (Pathway 3 in Figure 1A). The relative significance retention rate shows attenuated decrements with increasing matching ratios in MIMIC-IV compared to HiRID and eICU. LMEM coefficients mirror those derived from the LMEM analysis of matched training cohorts. The same analysis was also conducted to compare original and undersampled cohorts, in order to rule out the possibility that the above results are due solely to the reduced number of controls. Results in the appendix (Figure 7 and Figure 8) indicate that the absolute count of retained significant features in original versus undersampled training cohorts undergoes minimal reduction at 1:10 and 1:5 matching ratios. At the 1:2 matching ratio, dataset-specific patterns emerge: MIMIC-IV and, to a slightly lesser extent, eICU preserve nearly all significant features, whereas HiRID exhibits a substantial decrease. LMEM coefficients reveal negligible variation between original and undersampled training cohorts.

### 3.2 Sepsis Prediction Model Performance Declines After Case-Control Matching

The hyperparameter spaces utilized for Bayesian Optimization and their optimal values across different models and datasets are detailed in Table 6. XGBoost emerged as the slightly superior model across all three datasets when trained and tested with a one-hour data window and six-hour prediction horizon (see Methods section about windowing). The model achieved its highest performance on the HiRID dataset with an AUROC of 0.771 and nAUPRC of 3.400, followed by MIMIC-IV with an AUROC of 0.783 and nAUPRC of 3.152, while performance on eICU was lower with an AUROC of 0.715 and nAUPRC of 2.556 (Figure 5A). A consistent pattern emerged showing reduced model performance across all model architectures when trained and tested on the eICU dataset. Although the nAUPRC values demonstrated a 2-3.4-fold improvement over baseline predictive performance, the models still generated a number of false positive predictions, indicating room for further optimization.

We evaluated and compared model performance between undersampled (Pathway 1 in Figure 1A, case-control aligned (Pathway 2 in Figure 1A), case-control matched (Pathway 3 in Figure 1A), and original training sets without implementing model optimization strategies in Figure 5. All testing was conducted on the corresponding original testing sets to maintain consistency in evaluation. Our initial experiments utilized a one-hour observation window with a six hour prediction horizon (Figure 5B1), effectively requiring the models to predict labels for unmodified data rows. The models trained on original training sets demonstrated slightly lower AUROC and nAUPRC values compared to their Bayesian-optimized counterparts. Through ablation analysis, we discovered that excluding demographic features (sex, age, height, weight—shown as w/o demogr.) from both original training and testing sets did not negatively impact model performance. This finding further provides rationale for proceeding with our analysis of demographically case-control matched variants, as demographic features appeared non-essential for predictive accuracy. Substantial declines in model performance were observed across all datasets when training was conducted using case-control matched training sets (see Table 5 for training set sizes and sepsis prevalence). The severity of performance degradation was particularly striking, with AUROC values falling below 0.5, indicating performance worse than random chance, while nAUPRC values declined below 1, suggesting inferior performance compared to baseline predictions. These performance metrics demonstrated a clear inverse relationship with matching ratios, with the 1:2 matched cases and controls exhibiting the most pronounced deterioration in predictive capability. Interestingly, elevating sepsis prevalence through naive control undersampling significantly enhanced model performance, yielding maximum AUROC of 0.989 and maximum AUPRC of 41.09. A 1:10 matching ratio produced near-optimal AUROC metrics, with slight performance decrements observed at 1:5 and 1:2 ratios.Forest-based classifiers exhibited superior nAUPRC gains from undersampling (notably at 1:10 ratio) relative to gradient-boosted models. The same results are presented as AUPRC instead of nAUPRC in appendix (Figure 11). The appendix (Table 4) demonstrated comparable magnitudes of change in model performance metrics AUROC and AUPRC, with approximately half of the models showing slight improvements and half showing slight decrements (note that both AUROC and nAUPRC for these two matching methods are below/close to baseline performance and such small changes might not be meaningfully interpretable).

An interesting pattern emerged when comparing different model architectures: gradient-boosted models (LGBM and XGBoost) showed greater resilience in terms of AUROC values but demonstrated increased instability in nAUPRC metrics. Analysis of relative performance revealed a noteworthy cohort-specific effect, with the eICU cohort exhibiting more modest performance degradation compared to both HiRID and MIMIC-IV cohorts. This variable impact of case-control matching across cohorts suggests underlying differences in data structure and characteristics that influence model robustness. Examining different data and prediction window configurations revealed consistent patterns across our experiments. Performance peaked when models utilized six hours of input data with six hours of prediction horizon (Figure 5B2), demonstrating enhanced sepsis prediction capability under these conditions. The MIMIC-IV dataset yielded particularly decent results, with the LGBM model achieving the highest AUROC value of 0.817 and the XGBoost model reaching a peak nAUPRC value of 3.998. This may be due to the exceeding absolute numbers of significant group differences, as shown in the LMEM analysis (Figure 4B), paired with the data origin from a single hospital system. When we extended the prediction horizon to 10 hours beyond the data window while maintaining the six-hour data window, model performance aligned closely with our baseline models that used an one-hour data window and six-hour prediction horizon (Figure 5B3). A consistent trend emerged across all three windowing variants: models trained and tested on HiRID or MIMIC-IV consistently outperformed those utilizing the eICU dataset.

In summary, our experimental framework yielded model performance metrics that align with contemporary research findings across all investigated datasets, model algorithms, and temporal window configurations when using the original training set structure. However, when implementing absolute onset case-control matching with and without demographic characteristics to the training sets (both Pathway 2 and Pathway 3 in Figure 1A), we observed a substantial decrease in model performance across all experimental parameters. The matching ratio exhibited only a minor influence on performance outcomes, with an inverse correlation between model performance and matching ratio values. This suggests a paradoxical relationship in which absolute onset case-control matched training sets exhibit progressively higher sepsis prevalence, yet models fail to benefit from the reduced class imbalance and instead demonstrate difficulty in identifying predictive patterns.

## 4 Discussion

The goal of sepsis prediction algorithms is to identify patients at risk before the infection reaches a stage where treatment is less effective or major complications arise^52^. This remains a challenging task, as machine learning classification performance for sepsis demonstrates markedly lower accuracy compared to other ICU prediction tasks, such as mortality or acute kidney injury prognosis^53,54^. Enhanced methodological approaches are required to address the inherent challenges of pronounced class imbalance and optimize model performance. Clinical implementation necessitates robust model performance with an optimal balance between sensitivity and specificity to establish credibility among healthcare professionals.

### 4.1 The Matching Paradox in Sepsis Prediction

Many previous sepsis prediction studies have not explicitly used case-control matching, instead relying on observational datasets with imbalanced class distributions^30,37,55,56^. This methodological choice may contribute to the reported strong model performance, raising a critical paradox: could the absence of structured matching itself be influencing the observed predictive success? By addressing this issue, our study challenges the community to reconsider the best approach for developing safe and unbiased predictive models for sepsis prediction. We argue that understanding and mitigating potential biases arising from dataset construction choices is crucial for ensuring robust and generalizable clinical AI applications.

Our LMEM analysis revealed that case-control matching systematically reduced the number of features showing significant differentiation between sepsis and non-sepsis cases across all three ICU datasets. From an initial set of 35–43 significant features, matching at a 1:2 ratio retained only 24–29 significant features, representing a reduction of approximately 30%. While vital signs such as heart rate and blood pressure maintained significance across all matching variants, other traditionally important sepsis indicators displayed variable robustness. Notably, inflammatory markers like CRP exhibited reduced significance in MIMIC-IV and eICU matched variants, while lactate, a key indicator of tissue hypoperfusion, lost significance in the eICU variants. Moreover, we also found that performing “alignment” instead of full-fledged matching led to similar performance deteriorations. The higher performance observed in undersampled cohorts, compared to the original dataset, confirms that performance deterioration is attributable to the matching criteria rather than merely the reduced sample size. These findings suggest that matching procedures may inadvertently dilute crucial biological signals that serve as early sepsis indicators.

Our analysis revealed an inverse relationship between matching stringency and prediction model performance across all three harmonized ICU cohorts and the evaluated model algorithms, challenging conventional epidemiological assumptions about the benefits of matching for inference. The most severe performance degradation occurred with 1:2 matching, where AUROC values dropped below 0.5 and nAUPRC values fell below baseline levels. Notably, this performance drop was not simply due to data loss from matching—undersampled cohorts with the same ratio showed higher performance than the original unbalanced cohorts. Thus, the observed deterioration (across both alignment- and demographic-based matching) persisted despite increased sepsis prevalence in the matched datasets, indicating that matching-induced distortions in the feature space outweighed any potential benefits from class rebalancing. These findings were consistent across different temporal windows, with models showing optimal performance when using six hours of input data and minimal prediction windows.

Two potential mechanisms could explain these observations: (1) case-control matching may create artificial data distributions that obscure naturally occurring sepsis signals, particularly affecting gradient-boosted models, which showed varying stability in AUROC versus nAUPRC metrics; and (2) machine learning models developed without case-control matching may learn temporal patterns that, while predictive, are not directly related to sepsis pathophysiology. Our ablation analysis revealed that demographic features contributed minimally to prediction performance in unmatched datasets, suggesting these variables may not be critical for short-term sepsis prediction. However, recent research demonstrated that feature importance can vary significantly across prediction windows, with gender contributing 22% to 24-hour predictions but only 3% to 6-hour forecasts^57^. These findings highlight fundamental challenges in balancing epidemiological best practices with machine learning approaches for sepsis prediction, particularly when working across diverse clinical datasets and healthcare systems, and underscore the importance of considering prediction horizons when evaluating feature contributions.

### 4.2 Future Directions of Case-Control Matching

Several approaches could potentially mitigate the limitations observed with traditional demographic case-control matching in sepsis prediction. One promising direction involves expanding matching criteria beyond demographics to include baseline clinical characteristics. Implementing pre-onset trajectory matching, in which controls are matched based on vital sign trajectories during early ICU admission, could better account for temporal dynamics^58^. Similarly, creating strata using time-conditional percentiles of dynamic biomarkers such as lactate or white blood cell counts might improve matching precision. However, these approaches face practical constraints, especially related to data availability prior to sepsis onset; many patients meet sepsis criteria shortly after ICU admission, and blood-based clinical features often have delayed availability. Moreover, propensity score matching presents an alternative solution to address the dimensionality challenges inherent in traditional case-control matching^59^. This method calculates a composite score representing each patient’s probability of belonging to the case group based on multiple covariates, allowing for more efficient group balancing and enabling sophisticated matching approaches such as nearest neighbor or caliper matching. Although propensity score methods offer flexibility and reduce the need for arbitrary categorization of continuous variables, they generally require larger sample sizes for accurate score estimation.

Machine learning methods also offer new possibilities for dynamic matching optimization during model training. For instance, reinforcement learning-based feature masking could selectively retain features with high sepsis discrimination power while maintaining control over confounding variables. The DeepAISE framework illustrates this potential, achieving AUROCs between 0.87 and 0.90 for predictions made 4–12 h prior to sepsis onset through dynamic feature weighting across 24-hour windows^60^. These findings suggest that similar reinforcement architectures could optimize matching without compromising predictive capability. Additionally, generative adversarial networks (GANs) could synthesize matched controls, creating balanced datasets that preserve temporal covariate relationships while retaining critical sepsis signals. However, although synthetic controls offer advantages in data availability and near-perfect matching capabilities, they may introduce artificial patterns and lack the inherent biological variability found in real patient data. Consequently, the generalizability of models trained on such synthetic datasets could be limited.

### 4.3 Consequences of Dataset Harmonization

Recent advances in dataset harmonization across major publicly available ICU databases, such as HiRID, MIMIC-IV, and eICU, have substantially enhanced opportunities for multi-center validation of predictive models in intensive care research. However, while these harmonization processes standardize variable definitions and temporal resolutions, they may inadvertently compromise predictive power. Performance metrics from YAIB experiments^22^ demonstrate lower accuracy compared to state-of-the-art models trained on raw institutional data and similar publicly available datasets^2,56^. This discrepancy partly reflects YAIB’s focus beyond pure performance optimization, yet several specific harmonization-related impacts on data structure and granularity contribute to this performance gap. The standardization process can discard institution-specific predictive features embedded in local treatment protocols and documentation practices, effectively treating these latent prognostic signals as statistical noise. Furthermore, harmonized exclusion criteria can result in artificially homogeneous patient populations that lack the clinical heterogeneity necessary for developing robust, generalizable models. The temporal alignment of irregular clinical measurements to standardized time grids particularly affects the preservation of natural physiological trajectories crucial for dynamic prediction. This impact is especially pronounced for HiRID and eICU, which originally capture data at higher temporal resolutions than MIMIC-IV’s hourly intervals, leading to a significant loss of granular temporal information during harmonization. Nevertheless, these factors do not negate the fact that alignment and matching significantly diminished predictive power in comparison with unbalanced or original cohorts. It is also unwarranted to assume that even using non-harmonized versions of the datasets would have produced different results.

In terms of study limitations, several factors affect the interpretation and generalizability of our findings. Traditional case-control matching introduces intrinsic dimensionality challenges when matching characteristics are considered separately, particularly as the need for non-reusable matches increases the complexity of matching. While alternative approaches such as sex-based or random matching can help mitigate match availability issues, these simplified strategies may lead to suboptimal confounder control across the entire cohort. Our study’s cohort selection criteria and sepsis definition, following the YAIB workflow, resulted in notably lower sepsis prevalence compared to previous research using the same datasets. For example, a recent study reported a sepsis prevalence of 37% in HiRID^61^, while our analysis identified only 6.3%. The use of harmonized datasets further constrained feature availability. Although the SOFA score plays a central role in defining sepsis onset, critical components like vasopressor administration and Glasgow Coma Scale (GCS) scores were not available in these harmonized dataset versions. Additional clinician-guided harmonization efforts will be essential to expand the feature set, particularly regarding the medications and comorbidities^22^. Another limitation stems from our temporal definition of sepsis onset, which encompassed Sepsis-3 defined onset within a window spanning 48 h prior to and 24 h after the clinical suspicion of sepsis. This broad window complicates the ability to determine whether our models could predict sepsis earlier than the time of clinical suspicion. As noted in recent literature^52^, machine learning predictors should ideally identify at-risk patients before clinical suspicion arises—an ability our current study design cannot definitively demonstrate.

### 4.4 Conclusion

Our investigation highlights a fundamental challenge in sepsis prediction: models trained on unmatched data may capture patterns reflecting general ICU trajectories rather than sepsis-specific pathophysiology, potentially due to uncontrolled variations in length of stay and patient-specific monitoring durations. While case-control matching presents a promising strategy for improving model specificity and robustness, our results emphasize that matching methodologies must be carefully designed to preserve clinically relevant sepsis signals. The significant performance degradation observed with increasing matching stringency suggests that overly rigid approaches may inadvertently discard crucial predictive features. Striking the right balance between confounder control and preserving sepsis-specific patterns remains a critical challenge. Our findings underscore the need for more sophisticated preprocessing strategies in developing reliable machine learning models for sepsis prediction. Future research should focus on refining matching techniques to mitigate temporal bias while retaining pathophysiologically relevant signals—potentially by integrating dynamic matching constraints and clinical domain knowledge. Ultimately, by combining epidemiological rigor with advanced machine learning techniques, the field can move closer to delivering clinically actionable, generalizable sepsis detection tools that address the real-world complexities of critical care.

## Data Availability

All data produced are available on Physionet, and provided to authors based on credentialed access.

https://physionet.org/content/mimiciv/3.1/

https://physionet.org/content/hirid/1.1.1/

https://gitlab.ethz.ch/BMDSlab/publications/sepsis/case-control-matching-for-sepsis-prediction

## Acknowledgements

ChatGPT and Claude were used for code debugging. ChatGPT and DeepL were employed for assistance with writing to improve grammar, vocabulary, and fluency.

## Author contributions statement

Must include all authors, identified by initials, for example: A.A. conceived the experiment(s), A.A. and B.A. conducted the experiment(s), C.A. and D.A. analysed the results. All authors reviewed the manuscript.

## Appendix

### Supplemental Figures and Tables

**Figure 6.**
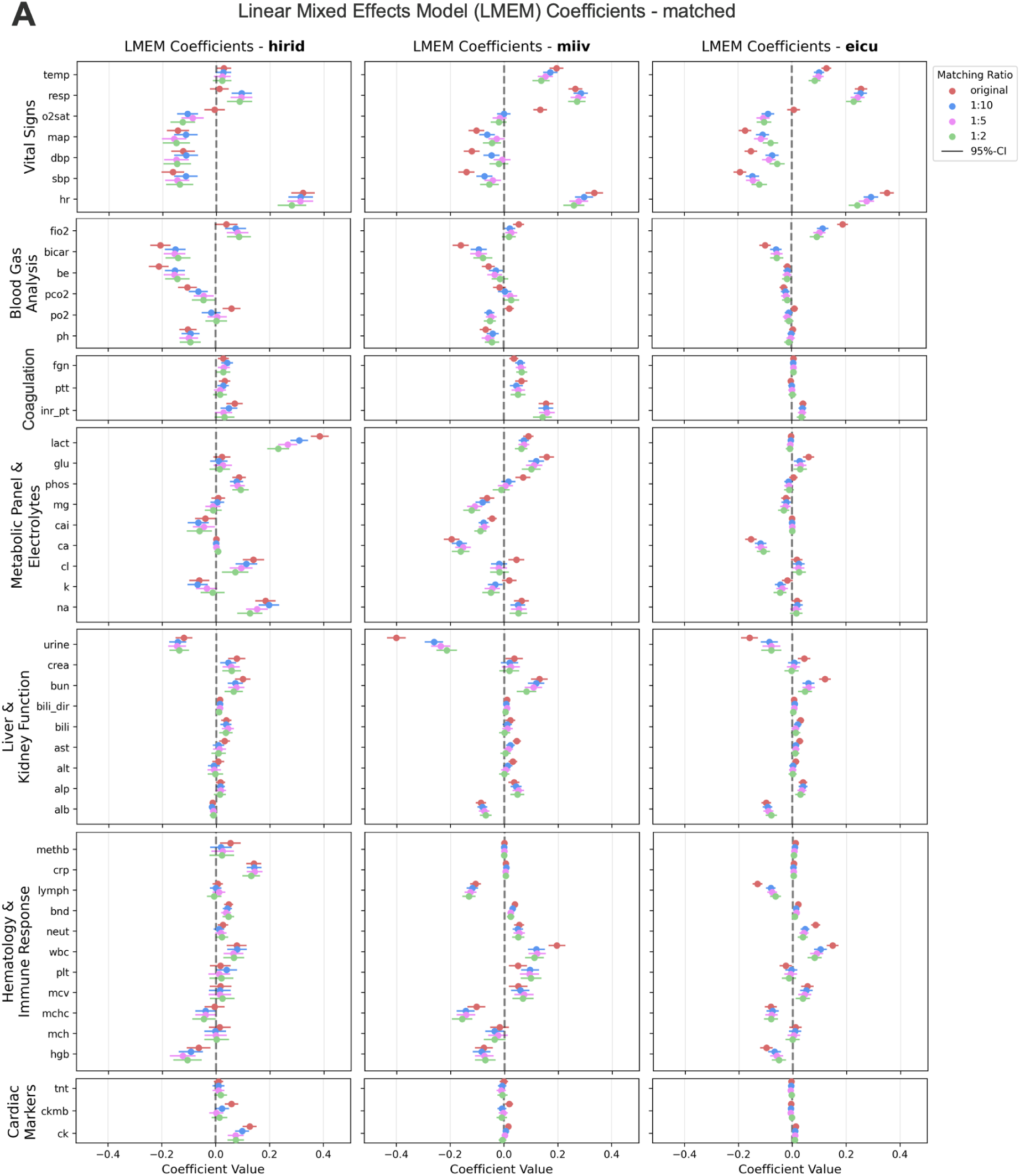
Absolute LMEM coefficients across datasets (original vs. matched). **A)** LMEM coefficients exhibit consistent patterns for dynamic features across all three datasets with higher magnitudes in original versus matched training cohorts, indicating attenuated discriminative capacity between sepsis and non-sepsis groups. Coefficients are plotted on the same scale for all grouped dynamic features and for all matched training cohort matching ratios. Positive coefficients indicate elevated feature values in the sepsis group relative to the non-sepsis group, whereas negative coefficients signify reduced values in sepsis cases. A coefficient of 1.0 denotes that the sepsis group exhibits, on average, a one-unit increase in the feature value compared to controls, after accounting for within-subject correlations via the mixed-effects model. All coefficients are presented with their respective 95% confidence intervals.

**Figure 7.**
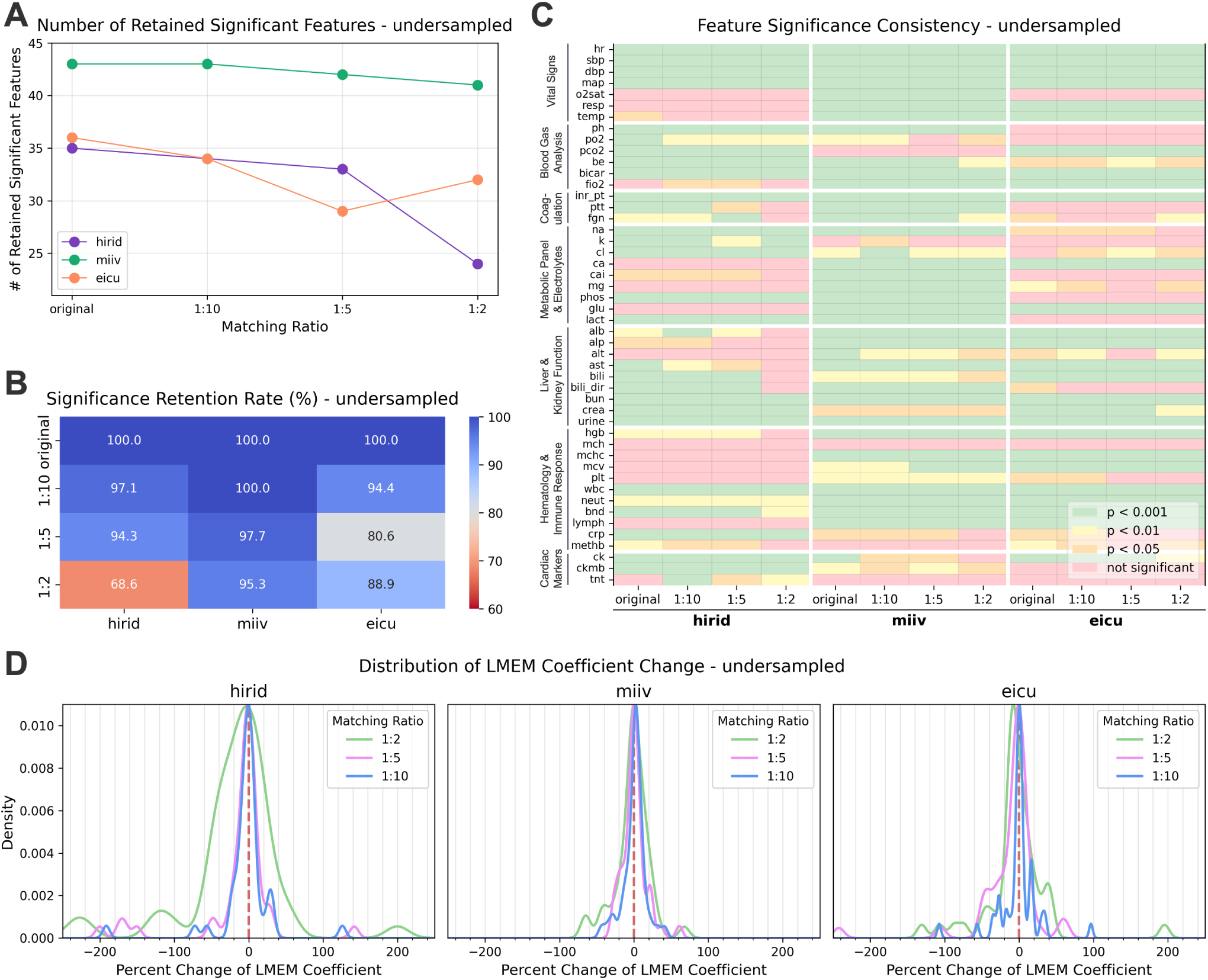
Evaluation of LMEM coefficient alterations and their statistical significance (original vs. undersampled). **A)** The absolute count of retained significant features in original versus undersampled training cohorts exhibits minimal reduction at 1:10 and 1:5 matching ratios. At 1:2 matching ratios, dataset-specific patterns emerge: MIMIC-IV and, to a slightly lesser extent, eICU preserve nearly all significant features, whereas HiRID demonstrates a substantial decrease. **B)** The relative significance retention rate similarly demonstrates dataset-specific patterns, with MIMIC-IV maintaining high preservation and HiRID exhibiting diminished retention at the 1:2 matching ratio. **C)** Analysis of significance levels reveals distinct heterogeneity in feature significance profiles across datasets. **D)** The percent change distribution in LMEM coefficients clusters around the original baseline with predominantly minor deviations, suggesting preserved discriminative capacity of collective dynamic features for sepsis versus non-sepsis cohort differentiation.

**Figure 8.**
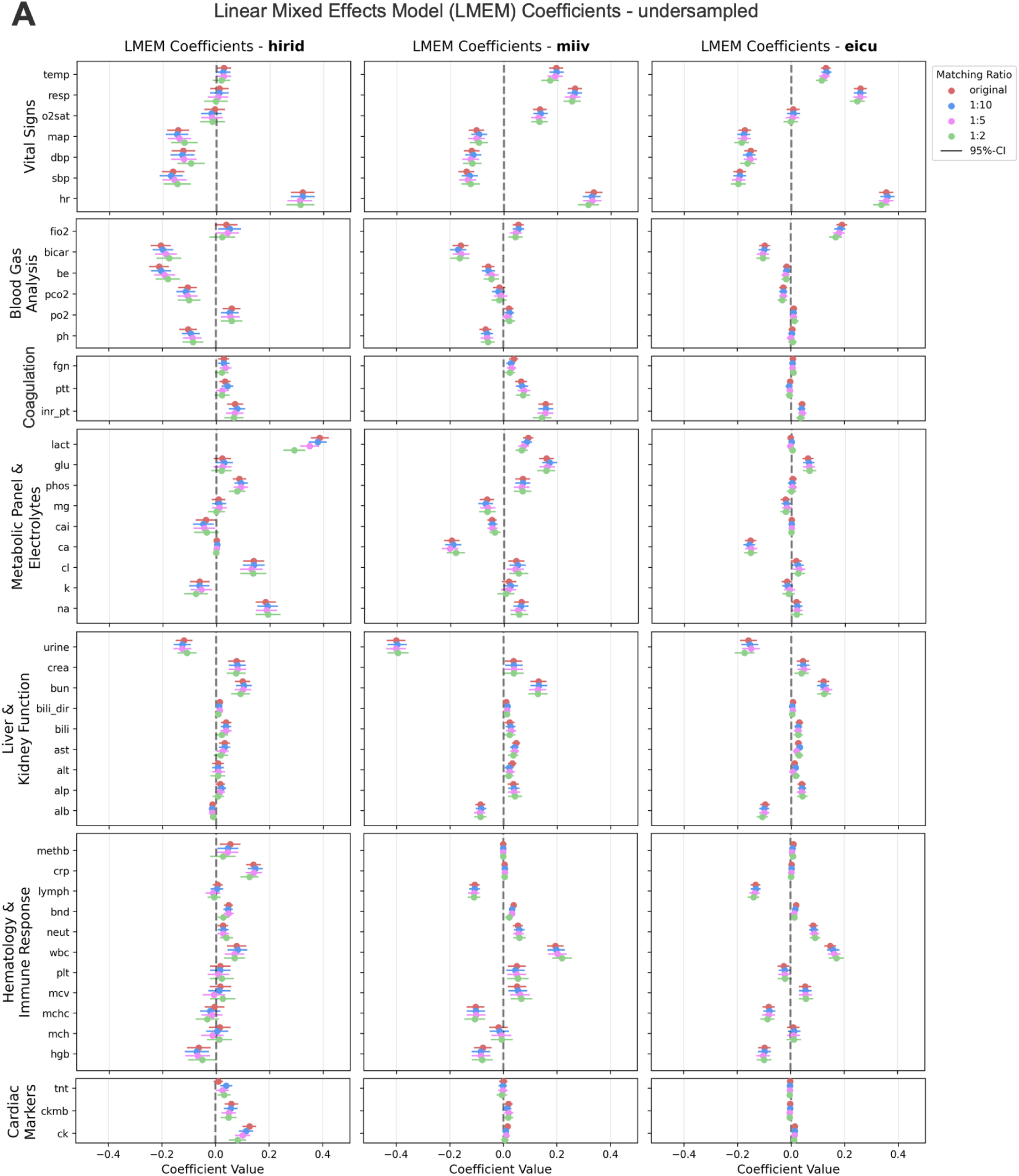
Absolute LMEM coefficients across datasets (original vs. undersampled). **A)** LMEM coefficients demonstrate negligible variation between original and undersampled training cohorts. Coefficients are plotted on the same scale for all grouped dynamic features and for all undersampled training cohort matching ratios. Positive coefficients indicate elevated feature values in the sepsis group relative to the non-sepsis group, whereas negative coefficients signify reduced values in sepsis cases. A coefficient of 1.0 denotes that the sepsis group exhibits, on average, a one-unit increase in the feature value compared to controls, after accounting for within-subject correlations via the mixed-effects model. All coefficients are presented with their respective 95% confidence intervals.

**Figure 9.**
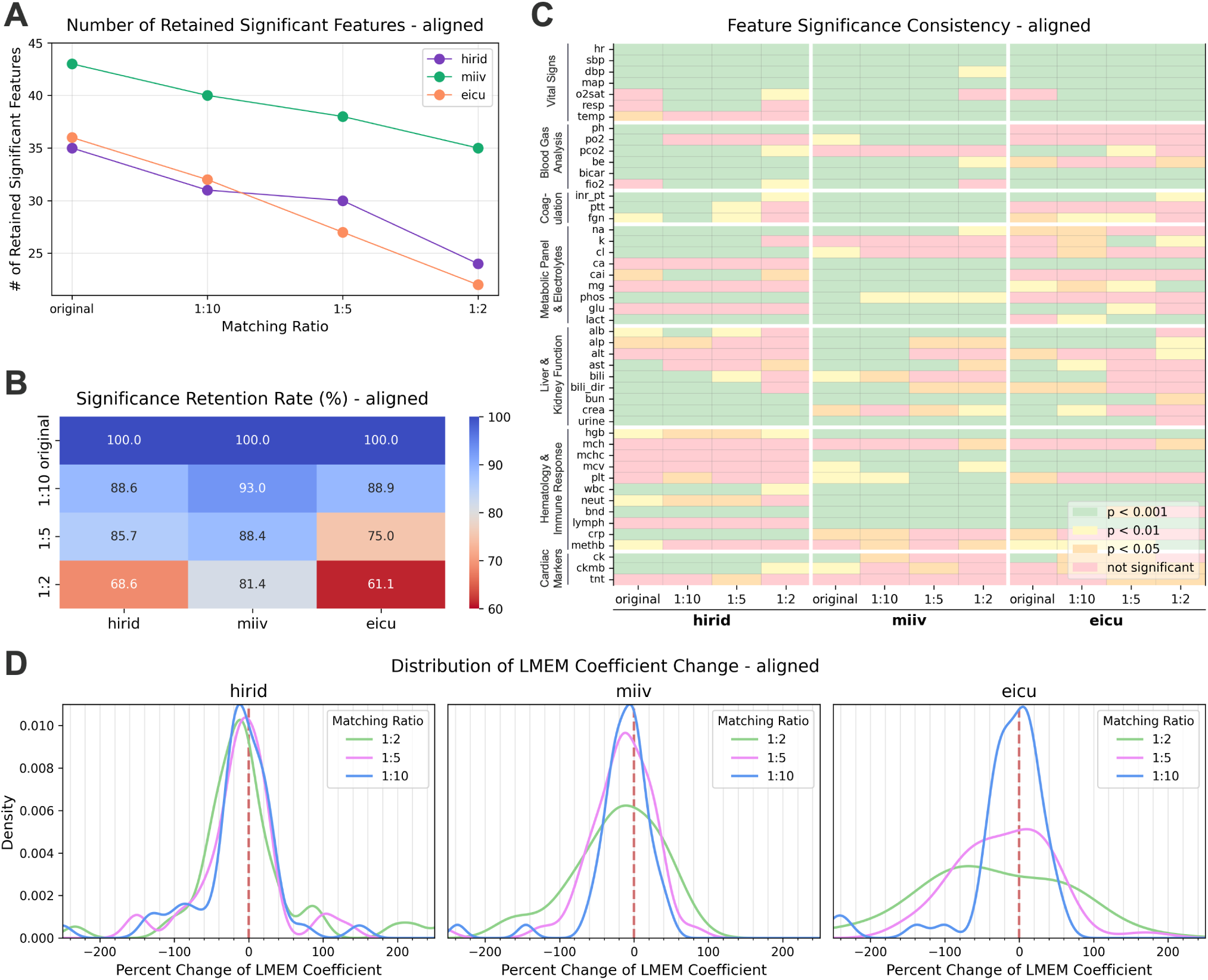
Evaluation of LMEM coefficient alterations and their statistical significance (original vs. aligned). **A)** The absolute number of retained significant features in aligned training cohorts compared to original training sets demonstrates an inverse relationship with increasing matching ratio. This relationship is comparable to results from matched training cohorts. The relative significance retention rate demonstrates attenuated decrements with increasing matching ratios in MIMIC-IV compared to HiRID and eICU. **C)** Analysis of significance levels reveals distinct heterogeneity in feature significance profiles across datasets. **D)** The distribution shift toward negative percent changes in LMEM coefficients indicates reduced discriminative capacity of the collective dynamic features for differentiating between sepsis and non-sepsis cohorts. Percent change values exhibited greater dispersion for eICU matching ratios of 1:5 and 1:2.

**Figure 10.**
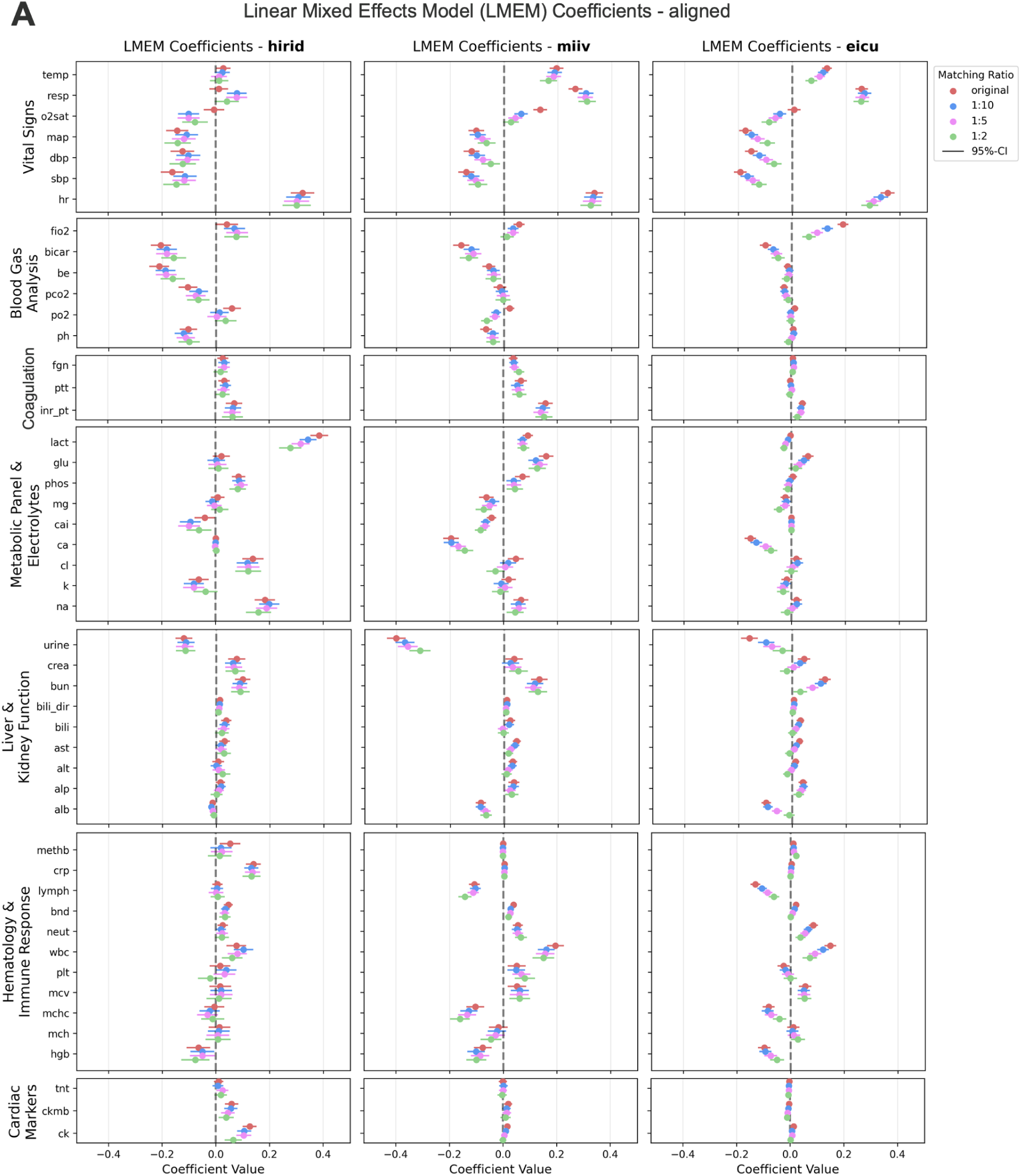
Absolute LMEM coefficients across datasets (original vs. aligned). **A)** LMEM coefficients parallel those derived from LMEM analysis of matched training cohorts. Coefficients are plotted on the same scale for all grouped dynamic features and for all aligned training cohort matching ratios. Positive coefficients indicate elevated feature values in the sepsis group relative to the non-sepsis group, whereas negative coefficients signify reduced values in sepsis cases. A coefficient of 1.0 denotes that the sepsis group exhibits, on average, a one-unit increase in the feature value compared to controls, after accounting for within-subject correlations via the mixed-effects model. All coefficients are presented with their respective 95% confidence intervals.

**Table 3.**
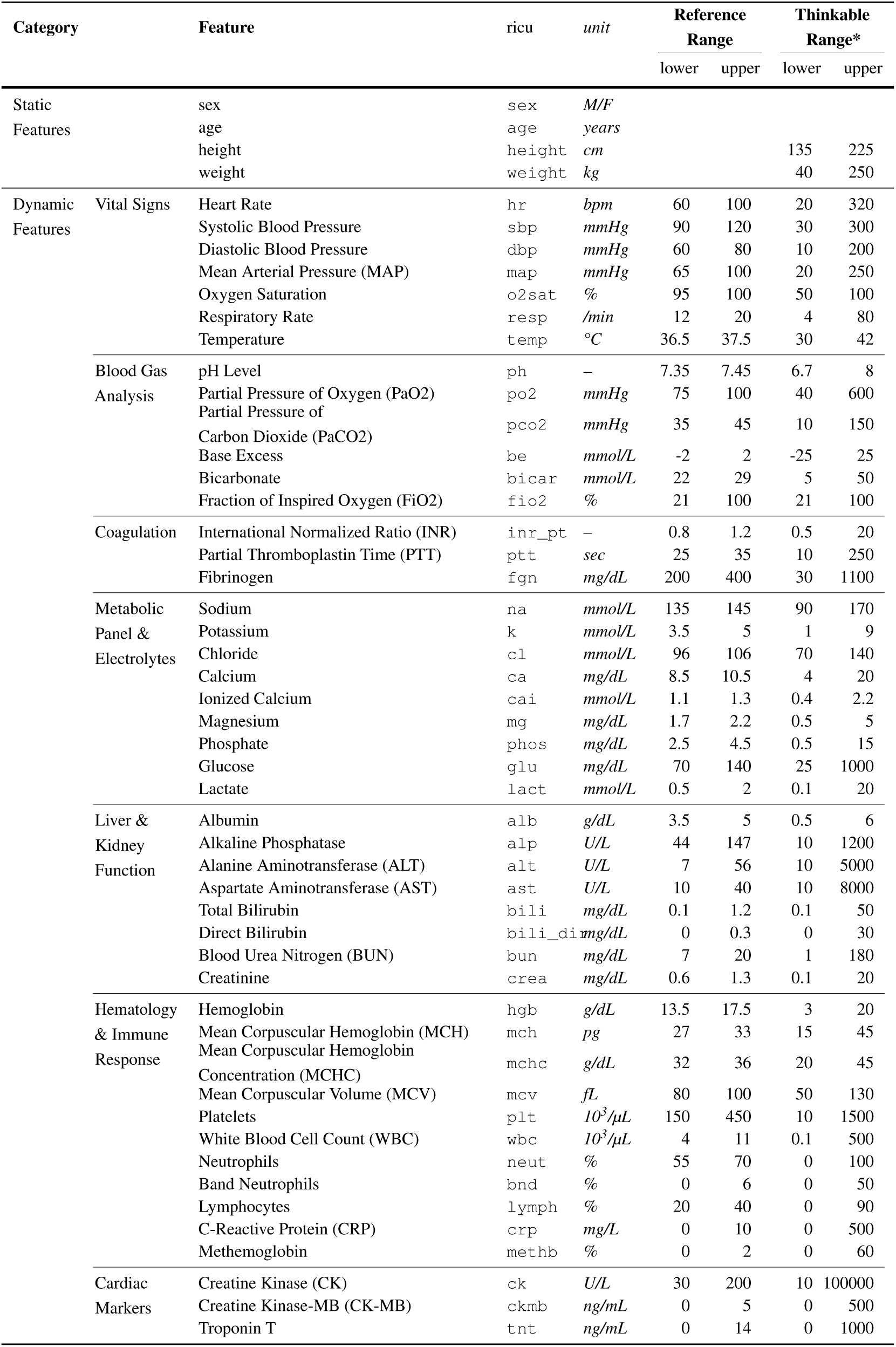
Overview of features in harmonized datasets. Clinical concepts were extracted using `ricu`. Reference and thinkable ranges were defined after harmonization to facilitate data interpretation and cleaning. *Ranges were defined with generous boundaries and with respect to the circumstances of ICU stays. Outliers that were under or over the limits by more than 5% of the thinkable range were replaced with NaNs.

**Figure 11.**
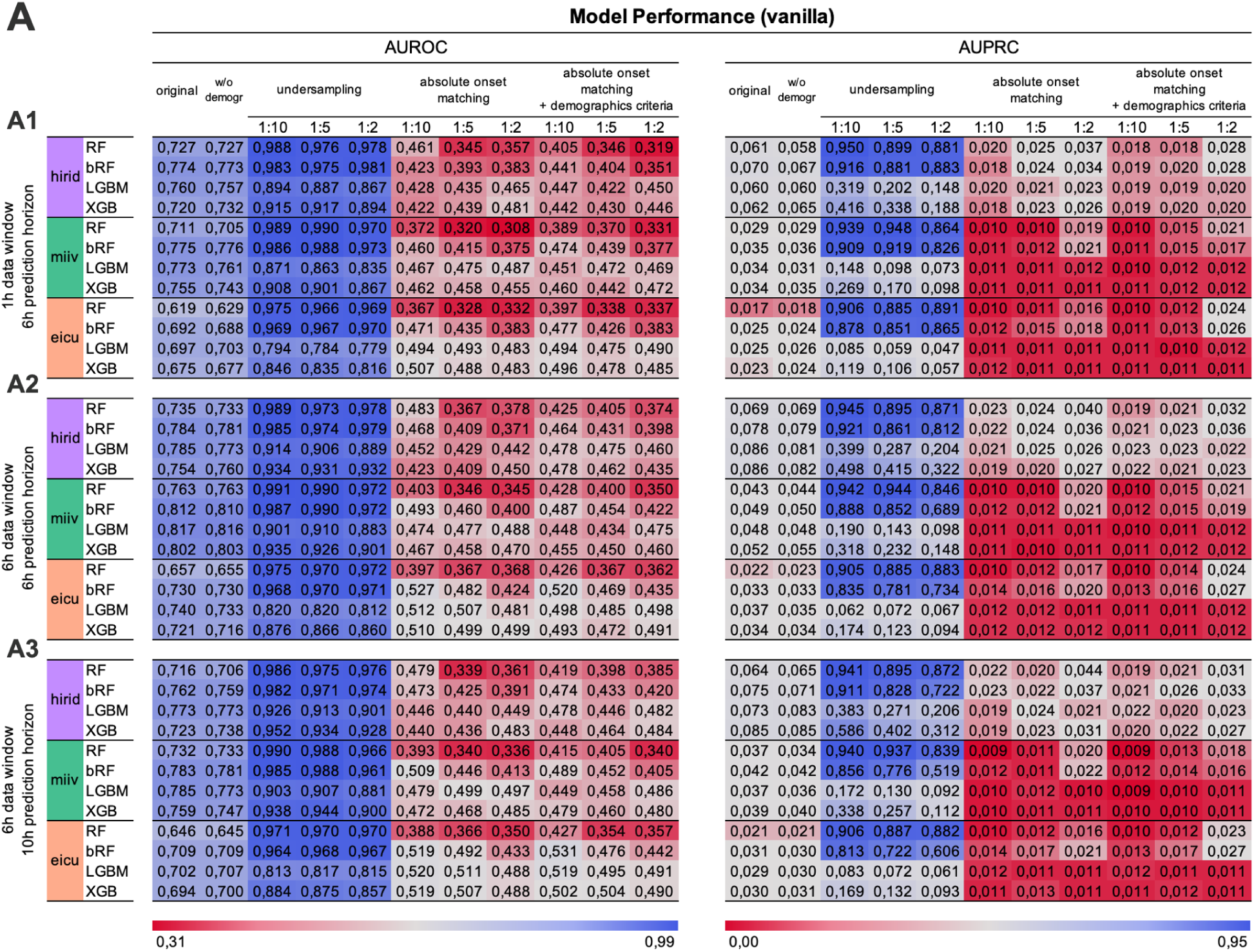
Performance of machine learning models. **A)** Comparative analysis of non-optimized (vanilla) machine learning models trained on specified training datasets (original, demographics-excluded, as well as undersampled, aligned, and matched at 1:10/1:5/1:2 ratios each) and evaluated on the original test cohort. Left panels display AUROC metrics while right panels present AUPRC metrics. Note that AUPRC values cannot be directly compared due to varying positive-class prevalence and therefore baseline performance across training cohorts (Table 5, Figure 12). This experimental framework was implemented across three distinct temporal configurations: **(A1)** 1-hour data window with 0-hour prediction window, **(A2)** 6-hour data window with 0-hour prediction window, and **(A3)** 6-hour data window with 4-hour prediction window. Models trained on undersampled cohorts demonstrate substantial performance metric improvements, while those trained on absolute-onset matched cohorts—with or without demographic criteria—exhibit performance metrics below baseline levels.

**Table 4.**
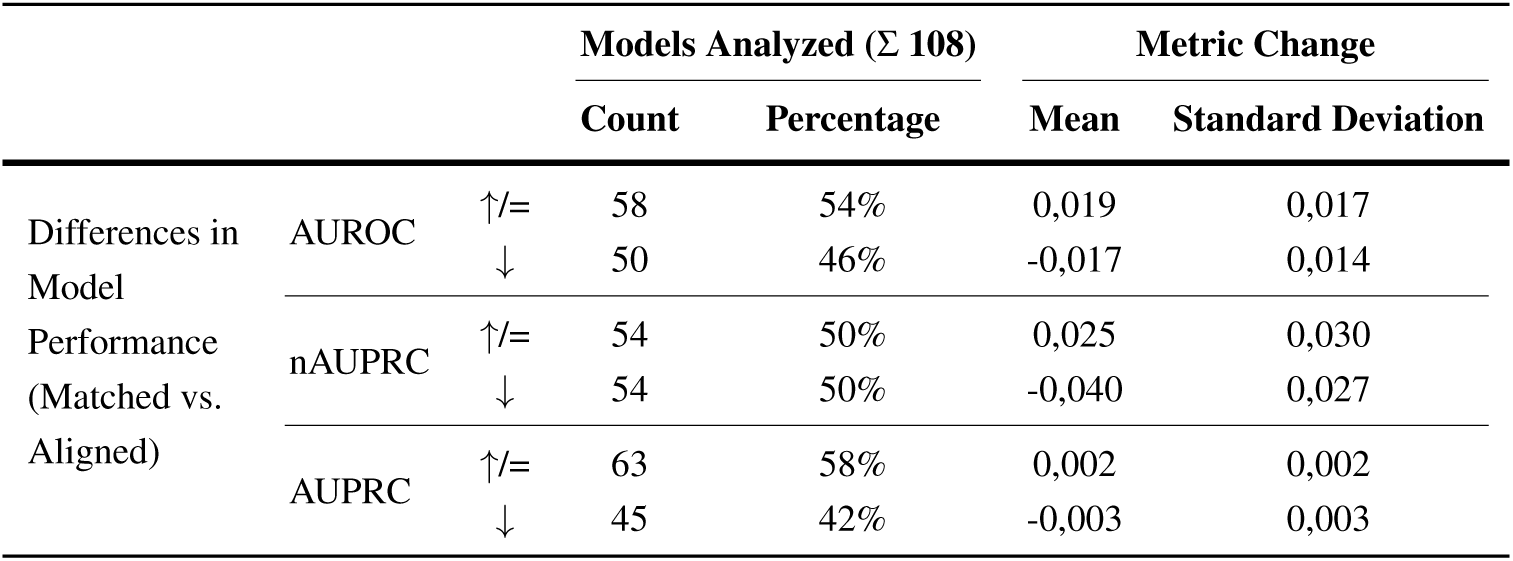
Differences in model performance metrics between matched and aligned cohorts. Arrows indicate performance improvement (↑), no change (=), or degradation (↓) of models trained on matched training sets (across all matching ratios) when compared to models trained on aligned training sets. A comprehensive evaluation was conducted across 108 distinct models, comprising a factorial design of 3 datasets, 4 model algorithms, 3 data-prediction-window configurations, and 3 matching ratios. The comparative analysis of matched versus aligned sampling methods reveals that the incorporation of additional demographic matching criteria produces minimal impact on model performance. The metrics AUROC, nAUPRC, and AUPRC demonstrate comparable magnitudes of change, with approximately half of the models showing slight improvements and half showing slight decrements in performance across each metric.

**Figure 12.**
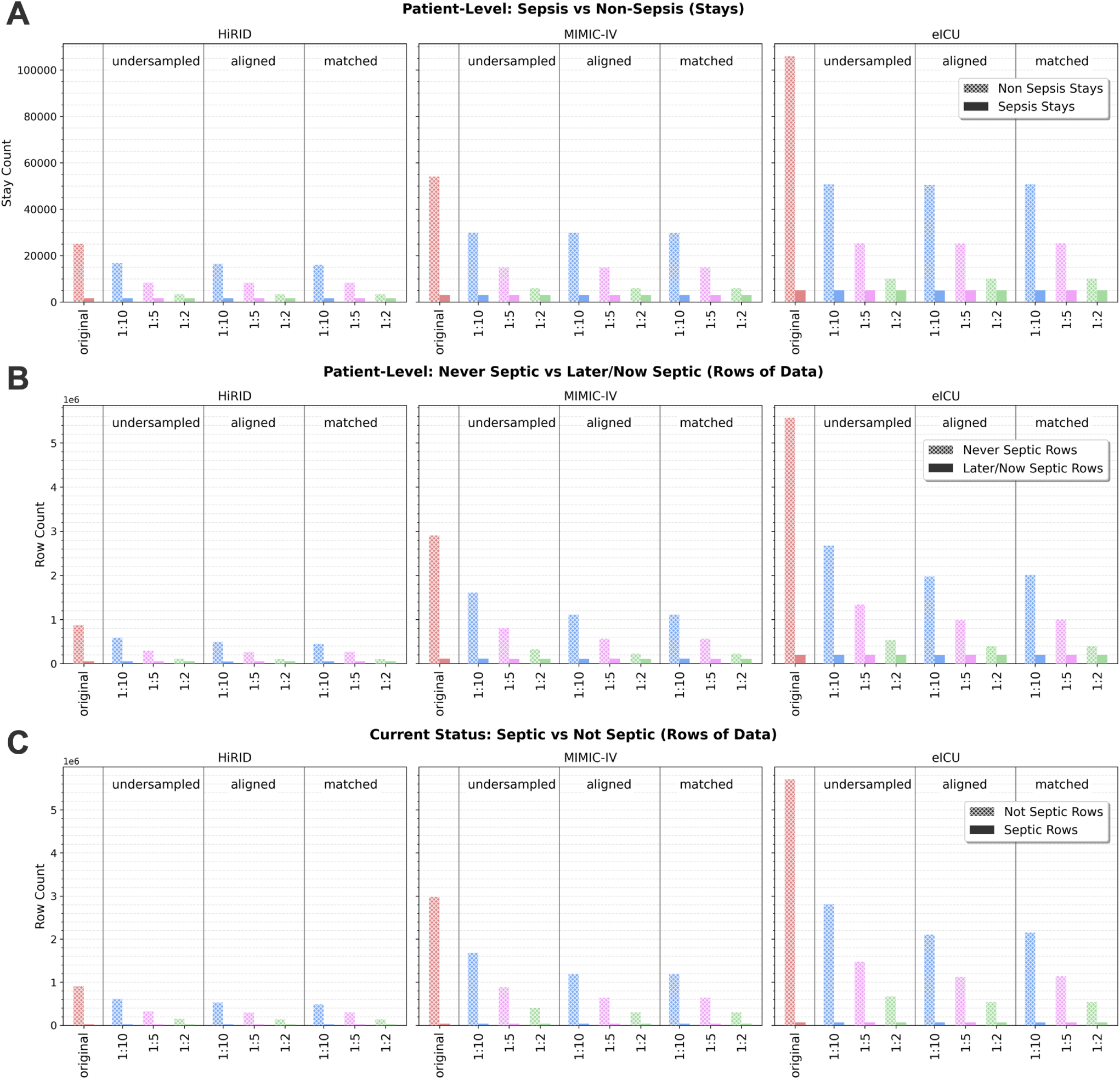
Overview of training set sizes and sepsis case and label prevalences. Implementation of three case-control matching ratios (1:10, 1:5, and 1:2) yields progressively reduced training cohorts with inversely increasing sepsis prevalence. Numerical data for these distributions are presented in Table 5. **A)** Number of stays in the sepsis and non-sepsis groups across different datasets (HiRID, MIMIC-IV, eICU), matching methods (undersampled, aligned, matched) and matching ratios (1:10, 1:5, 1:2). **B)** Combined number of rows (i.e. hours of data) of sepsis (later or now septic) and non-sepsis (never septic) stays. **B)** Number of rows (i.e. hours of data) with positive sepsis label (currently septic) and negative sepsis label (currently not septic).

**Table 5.** Summary of training and testing set characteristics across different matching methods and ratios. Machine learning analyses were conducted using the original dataset versions for both model training and evaluation, whereas the undersampled, aligned and matched dataset variants were exclusively utilized for model training. All models, regardless of training dataset, were subsequently evaluated on the original test sets to maintain external validity and enable assessment under clinically representative conditions. The total number of sepsis cases can vary slightly because of randomized splitting into

| Dataset | Matching Method | Variant | Set | Total Stays | Non-Sepsis Cases | Sepsis Cases (Prevalence) |
| --- | --- | --- | --- | --- | --- | --- |
| HiRID | original |  | train | 26,728 | 25,040 (93.7%) | 1,688 (6.3%) |
|  |  |  | test | 2,970 | 2,800 (94.3%) | 170 (5.7%) |
|  | undersampled | 10 | train | 18,394 | 16,726 (90.9%) | 1,668 (9.1%) |
|  |  | 5 | train | 10,023 | 8,354 (83.3%) | 1,679 (16.7%) |
|  |  | 2 | train | 5,016 | 3,343 (66.6%) | 1,673 (33.4%) |
|  | aligned | 10 | train | 18,394 | 16,726 (90.9%) | 1,668 (9.1%) |
|  |  | 5 | train | 10,011 | 8,350 (83.2%) | 1,668 (16.8%) |
|  |  | 2 | train | 5,016 | 3,343 (66.6%) | 1,673 (33.4%) |
|  | matched | 10 | train | 17,681 | 16,013 (90.6%) | 1,668 (9.4%) |
|  |  | 5 | train | 10,028 | 8,354 (83.3%) | 1,674 (16.7%) |
|  |  | 2 | train | 5,069 | 3,395 (67.0%) | 1,674 (33.0%) |
| MIMIC-IV | original |  | train | 57,082 | 54,058 (94.7%) | 3,024 (5.3%) |
|  |  |  | test | 6,343 | 6,046 (95.3%) | 297 (4.7%) |
|  | undersampled | 10 | train | 32,877 | 29,874 (90.9%) | 3,003 (9.1%) |
|  |  | 5 | train | 17,933 | 14,933 (83.3%) | 3,000 (16.7%) |
|  |  | 2 | train | 8,966 | 5,969 (66.6%) | 2,997 (33.4%) |
|  | aligned | 10 | train | 32,751 | 29,765 (90.9%) | 2,986 (9.1%) |
|  |  | 5 | train | 17,933 | 14,934 (83.3%) | 2,999 (16.7%) |
|  |  | 2 | train | 8,965 | 5,969 (66.6%) | 2,996 (33.4%) |
|  | matched | 10 | train | 32,734 | 29,737 (90.8%) | 2,998 (9.2%) |
|  |  | 5 | train | 17,933 | 14,946 (83.3%) | 2,987 (16.7%) |
|  |  | 2 | train | 8,966 | 5,969 (66.6%) | 2,997 (33.4%) |
| eICU | original |  | train | 111,071 | 105,988 (95.4%) | 5,083 (4.6%) |
|  |  |  | test | 12,342 | 11,787 (95.5%) | 555 (4.5%) |
|  | undersampled | 10 | train | 55,816 | 50,739 (90.9%) | 5,077 (9.1%) |
|  |  | 5 | train | 30,445 | 25,363 (83.3%) | 5,082 (16.7%) |
|  |  | 2 | train | 15,222 | 10,143 (66.6%) | 5,079 (33.4%) |
|  | aligned | 10 | train | 55,539 | 50,522 (91.0%) | 5,017 (9.0%) |
|  |  | 5 | train | 30,522 | 25,341 (83.0%) | 5,181 (17.0%) |
|  |  | 2 | train | 15,222 | 10,141 (66.6%) | 5,081 (33.4%) |
|  | matched | 10 | train | 55,816 | 50,734 (90.9%) | 5,082 (9.1%) |
|  |  | 5 | train | 30,445 | 25,363 (83.3%) | 5,082 (16.7%) |
|  |  | 2 | train | 15,222 | 10,155 (66.7%) | 5,067 (33.3%) |

**Table 5. Summary of training and testing set characteristics across different matching methods and ratios.**
| Dataset | Matching Method | Variant | Set | Total Rows | Group |  | Label |  |
| --- | --- | --- | --- | --- | --- | --- | --- | --- |
|  |  |  |  |  | Never-Septic | Later-/Now-Septic | Not Septic | Septic |
| HiRID | original |  | train | 930,608 | 877,612 (94.3%) | 52,996 (5.7%) | 908,712 (97.6%) | 21,896 (2.4%) |
|  |  |  | test | 102,473 | 96,175 (93.9%) | 6,298 (6.1%) | 100,267 (97.8%) | 2,206 (2.2%) |
|  | undersampled | 10 | train | 504,536 | 451,320 (89.5%) | 53,220 (10.5%) | 482,883 (95.7%) | 21,754 (4.3%) |
|  |  | 5 | train | 319,754 | 266,046 (83.2%) | 53,708 (16.8%) | 297,983 (93.2%) | 21,771 (6.8%) |
|  |  | 2 | train | 159,190 | 105,281 (66.1%) | 53,909 (33.9%) | 137,402 (86.3%) | 21,788 (13.7%) |
|  | aligned | 10 | train | 519,289 | 466,172 (89.8%) | 53,126 (10.2%) | 497,492 (95.8%) | 21,798 (4.2%) |
|  |  | 5 | train | 328,286 | 266,859 (81.3%) | 61,432 (18.7%) | 306,455 (93.3%) | 21,837 (6.7%) |
|  |  | 2 | train | 159,076 | 105,229 (66.1%) | 53,848 (33.9%) | 137,284 (86.3%) | 21,802 (13.7%) |
|  | matched | 10 | train | 504,519 | 451,264 (89.4%) | 53,255 (10.6%) | 482,883 (95.7%) | 21,636 (4.3%) |
|  |  | 5 | train | 319,754 | 266,046 (83.2%) | 53,708 (16.8%) | 298,047 (93.2%) | 21,714 (6.8%) |
|  |  | 2 | train | 159,404 | 105,563 (66.2%) | 53,891 (33.8%) | 137,598 (86.3%) | 21,836 (13.7%) |
| MIMIC-IV | original |  | train | 3,017,646 | 2,903,317 (96.2%) | 114,329 (3.8%) | 2,978,396 (98.7%) | 39,250 (1.3%) |
|  |  |  | test | 341,958 | 330,615 (96.7%) | 11,343 (3.3%) | 338,099 (98.9%) | 3,859 (1.1%) |
|  | undersampled | 10 | train | 1,722,288 | 1,608,255 (93.4%) | 114,037 (6.6%) | 1,683,395 (97.7%) | 38,896 (2.3%) |
|  |  | 5 | train | 978,358 | 865,528 (88.5%) | 112,832 (11.5%) | 939,470 (96.0%) | 38,893 (4.0%) |
|  |  | 2 | train | 489,048 | 376,253 (76.9%) | 112,795 (23.1%) | 450,127 (92.0%) | 38,942 (8.0%) |
|  | aligned | 10 | train | 1,272,709 | 1,152,570 (90.6%) | 119,530 (9.4%) | 1,234,222 (96.9%) | 38,528 (3.1%) |
|  |  | 5 | train | 697,878 | 584,053 (83.6%) | 114,440 (16.4%) | 659,128 (94.3%) | 39,570 (5.7%) |
|  |  | 2 | train | 337,942 | 225,240 (66.6%) | 113,034 (33.4%) | 299,211 (88.5%) | 38,795 (11.5%) |
|  | matched | 10 | train | 1,223,350 | 1,109,025 (90.7%) | 114,324 (9.3%) | 1,184,435 (96.8%) | 38,915 (3.2%) |
|  |  | 5 | train | 678,271 | 565,436 (83.4%) | 112,889 (16.6%) | 639,377 (94.3%) | 38,910 (5.7%) |
|  |  | 2 | train | 336,483 | 224,231 (66.6%) | 112,252 (33.4%) | 297,583 (88.4%) | 38,900 (11.6%) |
| eICU | original |  | train | 5,772,659 | 5,571,761 (96.5%) | 200,898 (3.5%) | 5,707,176 (98.9%) | 65,483 (1.1%) |
|  |  |  | test | 644,384 | 621,458 (96.4%) | 22,926 (3.6%) | 637,231 (98.9%) | 7,153 (1.1%) |
|  | undersampled | 10 | train | 2,575,364 | 2,370,363 (92.0%) | 205,001 (8.0%) | 2,512,803 (97.6%) | 62,561 (2.4%) |
|  |  | 5 | train | 1,530,332 | 1,330,332 (86.9%) | 200,001 (13.1%) | 1,467,619 (95.9%) | 62,713 (4.1%) |
|  |  | 2 | train | 737,419 | 535,312 (72.6%) | 202,204 (27.4%) | 671,967 (91.1%) | 65,432 (8.9%) |
|  | aligned | 10 | train | 1,609,570 | 1,375,572 (85.5%) | 198,653 (14.5%) | 1,514,923 (94.8%) | 84,647 (5.2%) |
|  |  | 5 | train | 920,273 | 721,472 (78.4%) | 200,503 (21.6%) | 838,823 (91.0%) | 83,064 (9.0%) |
|  |  | 2 | train | 602,885 | 400,605 (66.5%) | 202,349 (33.5%) | 537,348 (90.0%) | 65,484 (10.0%) |
|  | matched | 10 | train | 2,214,452 | 2,013,255 (90.9%) | 201,197 (9.1%) | 2,148,990 (97.0%) | 65,462 (3.0%) |
|  |  | 5 | train | 1,207,335 | 1,004,836 (83.2%) | 202,499 (16.8%) | 1,141,852 (94.6%) | 65,483 (5.4%) |
|  |  | 2 | train | 603,836 | 402,276 (66.6%) | 201,560 (33.4%) | 538,555 (89.2%) | 65,281 (10.8%) |

**Table 6.** Bayesian optimization hyperparameter spaces and best parameters. Hyperparameter ranges were defined for each machine learning model, with optimal parameters determined independently for each model-dataset pairing.

| Model | Hyperparameter | Range | Best Parameters |  |  |
| --- | --- | --- | --- | --- | --- |
|  |  |  | HiRID | MIMIC-IV | eICU |
| RF | n_estimators | 50-500 | 486 | 477 | 467 |
|  | max_depth | 3-30 | 19 | 16 | 13 |
|  | min_samples_split | 2-10 | 3 | 7 | 2 |
|  | min_samples_leaf | 1-4 | 2 | 3 | 2 |
| bRF | n_estimators | 50-500 | 409 | 415 | 400 |
|  | max_depth | 3-30 | 28 | 29 | 29 |
|  | min_samples_split | 2-10 | 9 | 9 | 9 |
| LGBM | n_estimators | 50-500 | 81 | 76 | 489 |
|  | learning_rate | 0.01-0.3 | 0.054 | 0.055 | 0.012 |
|  | max_depth | 3-30 | 6 | 7 | 15 |
|  | num_leaves | 20-100 | 92 | 89 | 62 |
| XGB | n_estimators | 50-500 | 188 | 179 | 478 |
|  | learning_rate | 0.01-0.3 | 0.053 | 0.092 | 0.028 |
|  | max_depth | 3-7 | 6 | 4 | 5 |
|  | min_child_weight | 1-5 | 4 | 3 | 2 |

## Code Availability

Scripts used to generate the results presented in this study are publicly available in a GitLab repository. The following Python scripts are included:

1. **Data Preparation**

- `preprocessing.py` This script utilizes harmonized datasets generated via the YAIB workflow (https://github.com/rvandewater/YAIB). It provides a flexible pipeline for specifying datasets (HiRID, MIIV, eICU), matching methods (original, undersampled, aligned, matched), matching ratios (1:2, 1:5, 1:10), and tolerances for strict demographics-based matching.
- `windowing.py` This script processes the preprocessed training and testing sets (output from `preprocessing.py`) to generate windowed datasets. Users can select preprocessed datasets by specifying the dataset (HiRID, MIIV, eICU), matching method (original, undersampled, aligned, matched), and matching ratio (1:2, 1:5, 1:10). Data and prediction window sizes are configurable (default combinations: 1/0, 6/0, 6/4).
2. **Linear Mixed Effects Models**

- `lmem.py` This script analyzes unwindowed preprocessed training sets (output from `preprocessing.py`) and trains univariate linear mixed effects models for each dynamic feature. Users can specify datasets (HiRID, MIIV, eICU) and matching methods (undersampled, aligned, matched), while original training sets are always included as a comparison. Output is provided as a text file report.
- `lmem_visualizations.py` This script converts the text file report (output from lmem.py) into a pandas DataFrame and generates visualizations.
3. **Machine Learning Models**

- `ml_vanilla.py` This script operates on preprocessed unwindowed (1-hour data window, 0-hour prediction window) or windowed training and testing sets (output from `preprocessing.py` and `windowing.py`). It offers a flexible framework for specifying datasets (HiRID, MIIV, eICU), machine learning algorithms (RF, bRF, LGBM, XGB), window configurations (default data-prediction-window combinations: 1/0, 6/0, 6/4), matching methods (original, undersampled, aligned, matched), and matching ratios (1:2, 1:5, 1:10). The script trains models and evaluates their performance.
- `ml_bayesianopt.py` This script utilizes preprocessed unwindowed (1-hour data window, 0-hour prediction window) training and testing sets (output from `preprocessing.py`). Users can specify datasets (HiRID, MIIV, eICU) and machine learning algorithms (RF, bRF, LGBM, XGB). The script applies Bayesian optimization to machine learning models, identifies optimal hyperparameters, and evaluates the resulting model performance.

